# Hidden Dependencies in Endometriosis Microarray Classifiers: Sample Reuse, Outcome-Informed Batch Correction, and Diagnostic Reassessment

**DOI:** 10.64898/2026.11.15.26368137

**Authors:** Elena Krikun, Abedalrhman Alkhateeb

**Affiliations:** Lakehead University, Thunder Bay, Ontario, Canada; Department of Computer Science, Lakehead University, Thunder Bay, Ontario, Canada

**Keywords:** endometriosis, GEO, Affymetrix CEL, exact sample reuse, data leakage, ComBat, scanner date, molecular classification, random forest, reproducibility, nested cross-validation

## Abstract

Distinct Gene Expression Omnibus (GEO) accessions do not necessarily represent independent transcriptomic samples. We investigated exact array reuse, access to test diagnoses during batch correction, and cohort and acquisition-date confounding in endometriosis classifiers. We compared decompressed CEL files using SHA-256, reconstructed a matrisome elastic-net workflow, changed test labels supplied to ComBat, and conducted 3,582 constrained training-label refits. We also evaluated the effect of removing matching training copies in a five-gene Huang-inspired random forest and performed a 324-probe nested internal benchmark within GSE51981. Of 37 test CEL files, 26 (70.3%) were identical to training arrays; all had been scanned in 2004–2005. Changing a test diagnosis supplied to ComBat moved the fixed classifier score toward the supplied label in all 37 interventions. In a deduplicated, batch-constrained comparison, median nominal balanced accuracy was 0.881 when ComBat received true test outcomes and 0.549 without them; the outcome-informed arm performed better in 1,193 of 1,194 matched refit comparisons. In the separate Huang-inspired reconstruction, removing each target’s matching training copy lowered pooled AUC from 1.000 to 0.323 (conditional 95% interval 0.105–0.571). Exploratory nested AUC was 0.733 for endometriosis versus other gynecological conditions, and date-held-out AUC in 2009 was 0.698. For endometriosis versus healthy controls, a date-only out-of-fold model achieved AUC 0.935, compared with 0.893 for the molecular model. Together, these findings show how exact array reuse, outcome-informed preprocessing, and acquisition-date structure can compromise the interpretation of nominally independent validation. They describe reconstructions and exploratory internal analyses, not corrected performance estimates for the published classifiers.

## 1 Introduction

Finding a molecular signature of endometriosis is not the same as showing that it can diagnose the disease in a new person. Gene expression reflects biology, but it also reflects when and how a specimen was collected, which tissue was profiled, which laboratory processed it, and how the resulting measurements were normalized. A classifier can learn some combination of these signals. Even a high test-set score is difficult to interpret if the purported test data have already appeared in training or if preprocessing requires the unknown diagnosis (Burney et al., 2007; Tamaresis et al., 2014; Cook et al., 2024; Kapoor and Narayanan, 2023).

Public repositories are especially vulnerable to subtle forms of reuse. The Gene Expression Omnibus (GEO) assigns distinct study and sample accession numbers, but those identifiers describe submissions, not necessarily unique biological specimens or measurements. An array can be deposited under two identifiers, analyzed again in a later study, or combined with other series. Legitimate reanalysis is a strength of open science; treating an already seen array as an independent test observation is a different matter. The distinction is particularly important when classifiers are trained on small datasets assembled from several sources.

Batch correction adds a second difficulty. ComBat estimates systematic differences between batches while allowing the analyst to specify biological covariates that should be preserved (Johnson et al., 2007). Using known diagnoses in a retrospective differential-expression analysis can be reasonable. In a diagnostic evaluation, however, the true diagnosis of a held-out patient is precisely the information the model is supposed to infer. Feeding that information to a joint preprocessing step changes the information available to the classifier, even if its fitted coefficients are held fixed. Such leakage should not be confused with ordinary overfitting: it occurs before a prediction is made. The potential for outcome-related batch adjustment to distort downstream inference and diagnostic evaluation has been described previously (Nygaard et al., 2016; Nieto et al., 2026); our contribution is an array-resolved intervention analysis of the present workflow.

A third difficulty concerns what an expression profile can inadvertently reveal about study origin. Cases and controls may have been sampled in different years, run in different laboratories, or represented by different tissue preparations. A model that appears to distinguish disease could also reflect those associated factors. The influence of technical batches on high-throughput measurements has long been recognized, as has the risk of overstating biological differences after covariate-aware batch correction (Leek et al., 2010; Nygaard et al., 2016). Cross-cohort portability must therefore be tested rather than assumed.

We began with the publicly released analysis of Cook, Wiggin and Fogg (Cook et al., 2024), which investigated the extracellular matrix (matrisome) in endometriosis and reported strong classifier performance. The availability of raw arrays and code made it possible to examine both sample identity and the flow of diagnostic labels through the analysis. As the audit developed, we extended it to other publications reusing the same GEO series, an independently implemented reconstruction inspired by the five immune-related genes of Huang et al. (2022), and a new internal benchmark in a single source cohort. These are *different* analyses: their balanced accuracies and ROC AUCs should not be read as before-and-after measurements of one original published model.

Our goal is to answer three practical questions. Where did the purportedly independent test measurements come from? Could preprocessing or exact sample reuse change predictions? And how much discrimination remains when we use more carefully contained, though still imperfect, internal evaluation? Figure 1 gives the overall workflow. Throughout, we distinguish verified array-level identity, conditional inferences about published sample membership, and features of models reconstructed from public descriptions.

**Figure 1:**
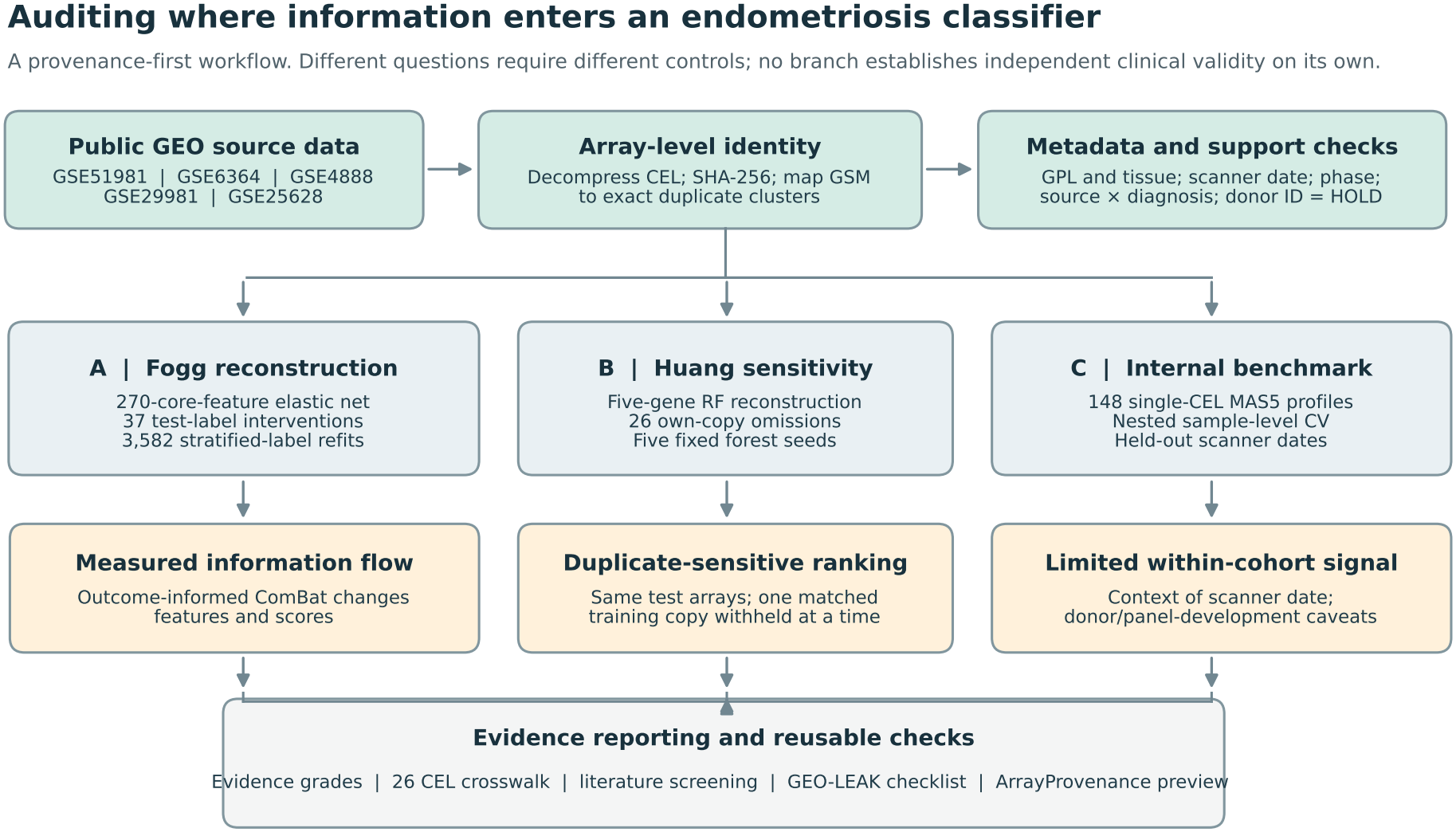
Study pipeline. Raw CEL identity and sample metadata were examined before any classifier analysis. Three branches address different threats: test-target access during batch correction, exact training-array reuse, and cohort/date-dependent generalization. The branches are not three estimates of the same clinical accuracy. The final output separates confirmed source evidence from unresolved donor or author-roster questions.

## 2 Materials and methods

### 2.1 Study design and the audit pipeline

This is a computational reproducibility study and a series of explicitly labeled sensitivity analyses. The units of observation differ across components. In the identity audit, the unit is a decompressed CEL file. In the literature review, it is a reported analytical use of a GEO series. In the model interventions, it is a fixed test measurement evaluated under a specified change to training or preprocessing. In the internal benchmark, the unit is a GEO sample, *not* a verified independent patient.

We organized the investigation into three quantitative branches after a common provenance review: (A) a reconstruction of the matrisome classifier and its outcome-informed ComBat processing, including label interventions and re-training nulls; (B) a fixed five-gene random-forest reconstruction with duplicate-copy omission; and (C) a single-CEL MAS5 internal benchmark that avoids the most direct test-label information paths. A parallel review documented scanner dates, tissue and platform differences, source-label association, and reported use of these arrays in later publications. The diagram and the detailed supplementary files make clear which questions were answered and which remained unresolved.

### 2.2 Raw-file provenance, partitions, and clinical metadata

The primary matrisome audit used GSE4888, GSE29981, GSE51981 and GSE6364. We reconstructed the published all-phase core-classifier layout of 179 training arrays (75 endometriosis cases and 104 controls) and 37 nominal test arrays from GSE6364 (21 cases and 16 controls). The original four-series archive inventory contained 232 CEL files, representing 206 unique decompressed contents. Our analysis of exact cross-series reuse additionally covered all 148 GSE51981 CEL files and 37 GSE6364 CEL files.

We decompressed individual CEL members before calculating SHA-256 hashes and linked matching hashes to GSE, GSM and file names. An exact match means that the underlying digital measurement is identical; it does not, by itself, reveal whether two different arrays came from the same donor. To create the deduplicated training layout, we removed only the 26 training arrays whose contents matched the fixed GSE6364 test set. This left 153 training arrays (56 cases and 97 controls) without changing the 37-array test set.

We recovered 148 scanner dates from the GSE51981 CEL headers using the affyio full-header reader, after an earlier text-only parser had failed on binary CEL headers. Dates were verified for all 148 source arrays, yielding 17 distinct days. Source-specific tissue, platform, phase, and clinical group information was reconciled from accession-level metadata and archived verified rosters. Within GSE51981, the clinical groups used here were 77 endometriosis (ENDO), 37 other gynecological conditions (OTHER), and 34 strict controls without pelvic pathology (NUPP). We did not treat the absence of a matching hash as proof of donor independence; a verified GSM-to-donor crosswalk was unavailable.

### 2.3 Matrisome reconstruction and controlled test-label interventions

We examined the publicly accessible code underlying the core-matrisome classification analysis of Cook et al. (2024), including the archived revision identified in the reproducibility files. The code forms a ComBat model matrix using the clinical condition covariate and performs batch adjustment jointly over the 216 training and nominal test arrays, with a reference batch. In the audited core reconstruction, the expression inputs resolve to 270 gene-level features after aggregating the microarray probes. This 270-feature implementation should not be conflated with the 258 core genes or 964 all-matrisome genes described in the publication: reconstruction of a downstream expression matrix does not certify identity to every author-trained coefficient.

We first checked the all-phase core-model confusion matrix. We then performed 37 one-at-a-time interventions: for each test array, we changed only the diagnosis value supplied to the ComBat model, not its measured expression or the true outcome used for scoring. Joint ComBat was re-estimated, and the already fitted reconstructed classifier scored the altered profile. We recorded whether expression features changed, whether the decision score moved toward the substituted diagnosis, and whether the predicted class crossed its threshold. Because ComBat was recalculated for all arrays, the experiment measures the response of the whole transformation, not a hypothetical isolated change in one feature.

In a separate set of 999 class-count-preserving assignments, we changed the *test labels supplied to ComBat* while holding the true scoring labels and fitted classifier fixed. Those distributions describe sensitivity of a particular retrospective pipeline to supplied outcomes. They are neither prospective accuracy estimates nor conventional sampling confidence intervals. We also examined two training layouts (179 and 153 arrays) under three processing options: joint ComBat with true test diagnoses, joint ComBat without an outcome covariate, and the prior per-series RMA representation without ComBat. Both ComBat alternatives still use the test expression distribution and are therefore transductive. Two previously fixed elastic-net hyperparameter settings were examined, and a post-hoc extension increased the solver iteration limit to resolve convergence warnings.

### 2.4 Retraining after constrained permutations of training diagnoses

The preceding interventions held a fitted classifier fixed. We next asked whether high nominal performance could persist when individual *training* diagnoses were reassigned and the classifier refitted from scratch. This extension was specified *after* the initial leakage findings, so it is reported as post-hoc rather than retrospectively described as preregistered.

For each eligible layout, we generated 199 seeded training-label assignments constrained to preserve the case and control totals within verified batches. For the original 179-array layout, a second assignment scheme also preserved the counts within batch-by-menstrual-phase strata. The corresponding deduplicated batch-by-phase scheme failed the prespecified support requirement and was not forced into the analysis. Each eligible assignment was fitted using RMA alone, joint ComBat without a diagnosis covariate, and joint ComBat with the true held-out diagnoses. We repeated the process with two fixed elastic-net parameter configurations. Critically,the outcome-informed arm still received the *true* test diagnoses at preprocessing time, even when the training diagnoses had been reassigned.

This produced 18 evaluable conditions (three layout/stratification designs, three preprocessing arms, two fixed model settings), each with 199 model refits: **3**,**582 fitted classifiers** in total. Of these, 2,388 used one of the two ComBat variants and 1,194 used RMA alone. Six contrasts paired an outcome-informed ComBat refit with the outcome-free ComBat refit trained under the same reassigned labels: 6 *×*199 = 1,194 matched comparisons. We assessed balanced accuracy (mean of sensitivity and specificity) against the unchanged 21-case/16-control test outcomes, and summarized empirical distributions and paired differences. Within-stratum label reassignment can leave source- or phase-associated information intact. Accordingly, these are controlled intervention distributions, not an exchangeable disease-null test or an estimate of true clinical performance inflation.

### 2.5 Published reuse, source composition, and scanner-date controls

We assembled a deliberately targeted literature index using GEO accession searches. The search returned 246 bibliographic records, 29 of which had indexed matches to both GSE51981 and GSE6364. Twenty-seven of these had accessible full text or methods examined, while two were reviewed using abstracts or version-level information. For each candidate we distinguished: (i) documented opposite train/test roles, (ii) pooled use without a claimed independent test, (iii) explicitly excluded series, and (iv) unknown exact sample selection. The search is not exhaustive and does not provide a valid denominator for the prevalence of leakage in all endometriosis studies.

We also compared the 324 fixed probe signals from independently processed individual CEL files across the three training studies, and contrasted differences between controls from different series with ENDO-versus-control differences within GSE51981. We examined the three original menstrual-phase tokens without combining them after observing the results. These probe-level comparisons describe expression structure; probe IDs must not be counted as independent biological biomarkers.

To investigate whether scanner-date sensitivity extended to previously reported expression markers, we prespecified ten genes from the published Huang and Chen signatures and summarized their MAS5 log_2_ contrasts on three supports: all eligible arrays; only dates containing both diagnoses without adjustment; and exactly that same restricted set with scanner-date fixed effects. The last two estimates deliberately use identical arrays, separating sample-support changes from the effect of covariate adjustment. We report conditional sample-bootstrap intervals, date-stratified permutation checks and Holm correction across 20 fixed-effect comparisons (Supplementary Section S11); this is a post-hoc association analysis, not a new diagnostic model.

Scanner-date controls used the same stored sample-level outer folds as the new molecular benchmark. Each date-only probability was constructed from the *training* portion of that fold, with smoothing for small counts; it was then applied to held-out samples. Year-only controls were calculated separately. We additionally summarized discrimination using only case–control pairs with the same exact scanner date. That within-date statistic uses fixed predictions and is not a new date-adjusted fitted model. A post-hoc 2009-only leave-one-date-out analysis was conducted only for the mixed-class ENDO-versus-OTHER setting, using five eligible scanner dates and nested training-fold-only model selection; 199 within-date label permutations served as a conditional sensitivity comparison.

### 2.6 A new annotation-derived internal expression benchmark

The new internal benchmark is distinct from either published classifier. A 275-gene core-matrisome source list (Naba et al., 2012) and the GPL570/GPL571 annotation files deterministically reproduce the 193-gene/324-exact-probe panel used here without consulting diagnostic labels. The probe-set identity was fixed before this benchmark. The exact annotation-based reconstruction demonstrates that the final panel can be obtained without outcome labels. As in any retrospective analysis, this does not by itself certify that all decisions about the feature-space definition were independent of earlier exploratory results (Ambroise and McLachlan, 2002).

We obtained one MAS5 signal vector from each of 148 GSE51981 CEL files; 118 were available from an earlier archived technical extraction and 30 additional CEL files were processed independently. We evaluated three fixed diagnostic tasks: ENDO versus all non-ENDO (77/71), ENDO versus OTHER (77/37), and ENDO versus NUPP (77/34). Standardization and logistic-regularization selection were performed within training portions of five outer and four inner folds. We preserved outer-fold prediction scores for all task-eligible records and compared with phase-only and intercept-frequency baselines. A predeclared 2009-only comparison used 46 ENDO and 31 OTHER arrays. We report sample-level out-of-fold ROC AUC, balanced accuracy, and Brier score, plus 1,000 bootstrap resamples of fixed out-of-fold scores. These intervals do not account for donor clustering, uncertainty from feature-rule development, or re-training every bootstrap sample.

### 2.7 Five-gene reconstruction inspired by Huang and own-copy omission

Huang et al. (2022) identified five immune-related genes (ACKR1, LMNB1, MFAP4, NMU, SEMA3C) and reported a published external AUC of 0.656 for a diagnostic signature. We did not recover that study’s fitted classifier or duplicate its original sva preprocessing. Instead, we constructed an explicitly separate five-gene, 1,000-tree random forest using independently extracted MAS5 signals: 37 arrays from GSE6364 and 22 from GSE25628 for training (59 total), followed by GSE51981 for evaluation. GSE25628 itself contains distinct eutopic, ectopic and control tissue types, another reason not to call this an exact reproduction of the original clinical evaluation.

For each of the 26 GSE51981 arrays with an identical training-side CEL, we refitted the fixed random-forest specification after omitting *only that array’s own exact training counterpart*. Each refit kept 58/59 training profiles and predicted the unchanged target. As a size- and class-matched control, we instead omitted a different training array of the same class from GSE6364 while keeping the exact target copy. The target-specific scores were pooled to compute a cross-fit AUC among the 26 repeated arrays; this is not the AUC of a single, externally validated forest. In a post-hoc model-randomness sensitivity check, we repeated the fixed experiment with five specified random seeds. A 5,000-resample class-stratified bootstrap of the fixed target scores provided conditional uncertainty intervals; it was not a full bootstrap of the complete training procedure.

### 2.8 Evidence grades and safeguards against overinterpretation

Exact CEL hashes demonstrate identity of a digital measurement but do not establish patient independence. Published study totals can imply conditional lower bounds on sample overlap, but cannot replace author-specific GSM rosters. Comparisons across preprocessing choices or different clinical control groups address different estimands and should not be interpreted as a direct numerical correction to published accuracy. Supplementary Section S11 documents the assayability and calendar overlap of ten published marker genes, but no new gene-level association claim is used to support our main classifier conclusions.

### 3 Results

### 3.1 One GEO accession was not one independent source of test arrays

We found 26 byte-identical CEL pairs between GSE51981 and GSE6364, corresponding to 26/37 (70.3%) nominal test arrays in the reconstructed core-classifier split. The repeated test arrays included 19 of 21 cases (90.5%) and seven of 16 controls (43.8%). The other 11 test records included only two cases and nine controls, making performance within this small subset highly imprecise. All 26 exact matches were recovered again from the complete source RAW archives, independently of expression normalization.

The date crosswalk revealed a striking structure: all 17 arrays scanned in GSE51981 in 2004 and all nine scanned in 2005 had identical GSE6364 counterparts. None of the 122 arrays scanned between 2009 and 2011 did. The 26 repeats comprised 19 ENDO and seven NUPP observations; no OTHER sample was an exact match. This links the repeated digital measurements to the earliest scanner-date strata without establishing why those measurements were archived more than once (Figure 2).

**Figure 2:**
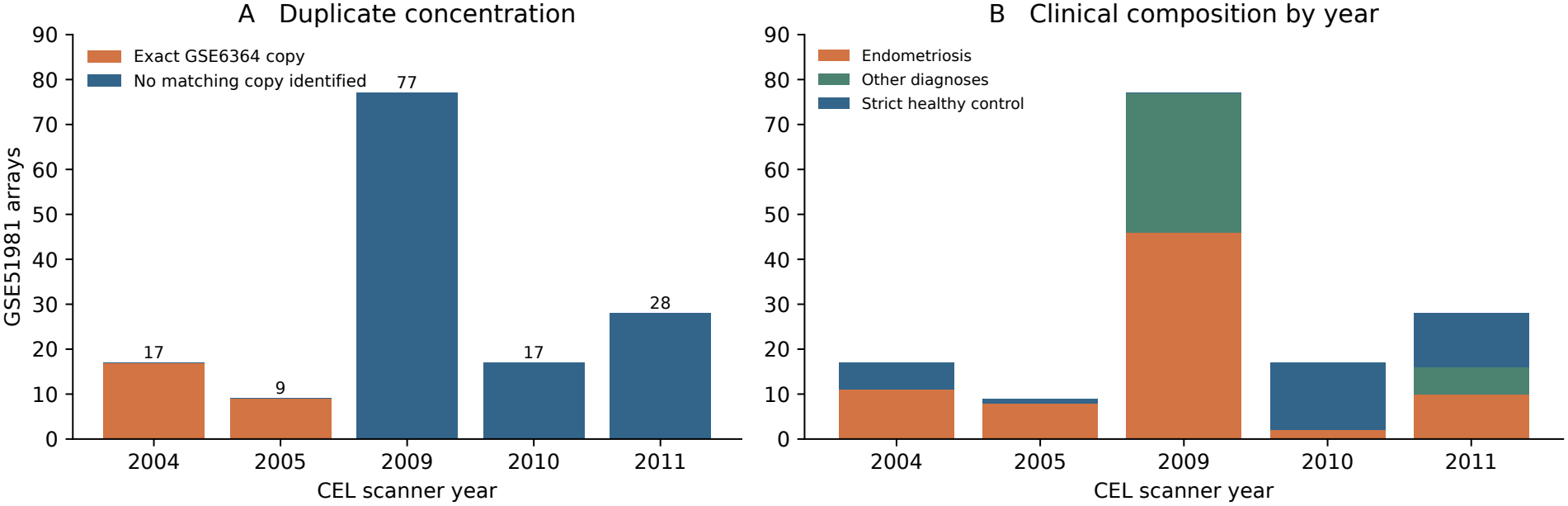
Raw-array reuse and scanner-year structure. Year-specific counts of GSE51981 CEL files with and without an exact GSE6364 counterpart, alongside the clinical group composition. Exact SHA-256 equality identifies reused digital measurements, not independent donors or a history of disclosure.

### 3.2 True test diagnoses changed the input of the reconstructed core classifier

Replaying the core matrisome analysis gave 19 true positives, two false negatives, 16 true negatives and no false positives on 37 nominal test arrays (balanced accuracy 0.9524). When we changed one test diagnosis *only in the ComBat input*, all 270 corrected features for that array changed. In all 37 one-at-a-time interventions, the fixed classifier score moved toward the substituted diagnosis; 16/37 interventions also switched the predicted class. This is a direct demonstration that the unknown test target could alter both features and predictions within the reconstructed pipeline. It does not establish that the original biological associations were incorrect.

Under 999 class-count-preserving assignments of supplied test labels, the median balanced accuracy of the fixed classifier fell to 0.7396 (empirical 5th–95th percentile range 0.6458–0.8192), compared with 0.9524 when the true diagnoses were supplied. The score used for evaluating each assignment was still the *real* diagnosis; only the label presented to ComBat had been changed. Thus these are preprocessing interventions, not a permutation test of disease prediction in a new cohort.

The converged 2-by-3 sensitivity analysis showed that removing 26 duplicate training arrays did not by itself remove high apparent accuracy while true test diagnoses remained available to ComBat (Table 1). With deduplicated training, balanced accuracy was 0.9524 for outcome-informed joint ComBat, 0.6592 for outcome-free joint ComBat, and 0.5000 for the previously processed RMA-only arm. The corresponding full-training values were 0.9524, 0.7857 and 0.6190. Neither alternative joint-ComBat approach is an independently deployable new-batch transformation because both see the target expression distribution.

**Table 1.** Balanced accuracy of the converged reconstructed core-matrisome classifier under different processing and training layouts.

| Processing used before classification | Original 179 train | Deduplicated 153 train |
|---|---|---|
| Joint ComBat with true test outcomes | 0.9524 | 0.9524 |
| Joint ComBat without outcomes | 0.7857 | 0.6592 |
| Per-series RMA without ComBat | 0.6190 | 0.5000 |

The same 37 test arrays were scored in every arm; true test outcomes were retained for evaluation. Both ComBat arms use test expression in harmonization. Training composition and feature transformations differ, so the numeric differences are not isolated causal estimates of leakage.

### 3.3 High apparent accuracy persisted after 3,582 training-label refits

The more stringent intervention was to *refit* classifiers after shuffling the individual training diagnoses within verified groups. We completed 3,582 fits across 18 evaluable conditions. One especially informative comparison used 153 deduplicated training arrays, training labels reassigned within batches, and the archived elastic-net configuration. Median nominal test balanced accuracy was **0.881** when ComBat still received the true test diagnoses, compared with **0.549** without a diagnosis covariate (199 refits for each arm). For the corresponding original 179-array training layout, the medians were 0.850 and 0.551; with batch-and-phase-constrained reassignment of 179 labels they were 0.857 and 0.558.

Across the six fixed matched ComBat comparisons, paired median balanced-accuracy advantages of the outcome-informed arm ranged from +0.260 to +0.317. It was higher in 1,193 of 1,194 replicate-matched comparisons. These repeated fits always used the *same 37 test arrays*, not 1,194 independent patient samples. The RMA-only branch assigned every test case to the control class in 988/1,194 fits, so apparent balanced accuracy near 0.5 there often reflected a degenerate all-control classifier rather than a distribution of random predictions. Figure 3 shows the complete outcome-dependent pattern; the 18-condition distributions and full paired table remain in the Supplement.

**Figure 3:**
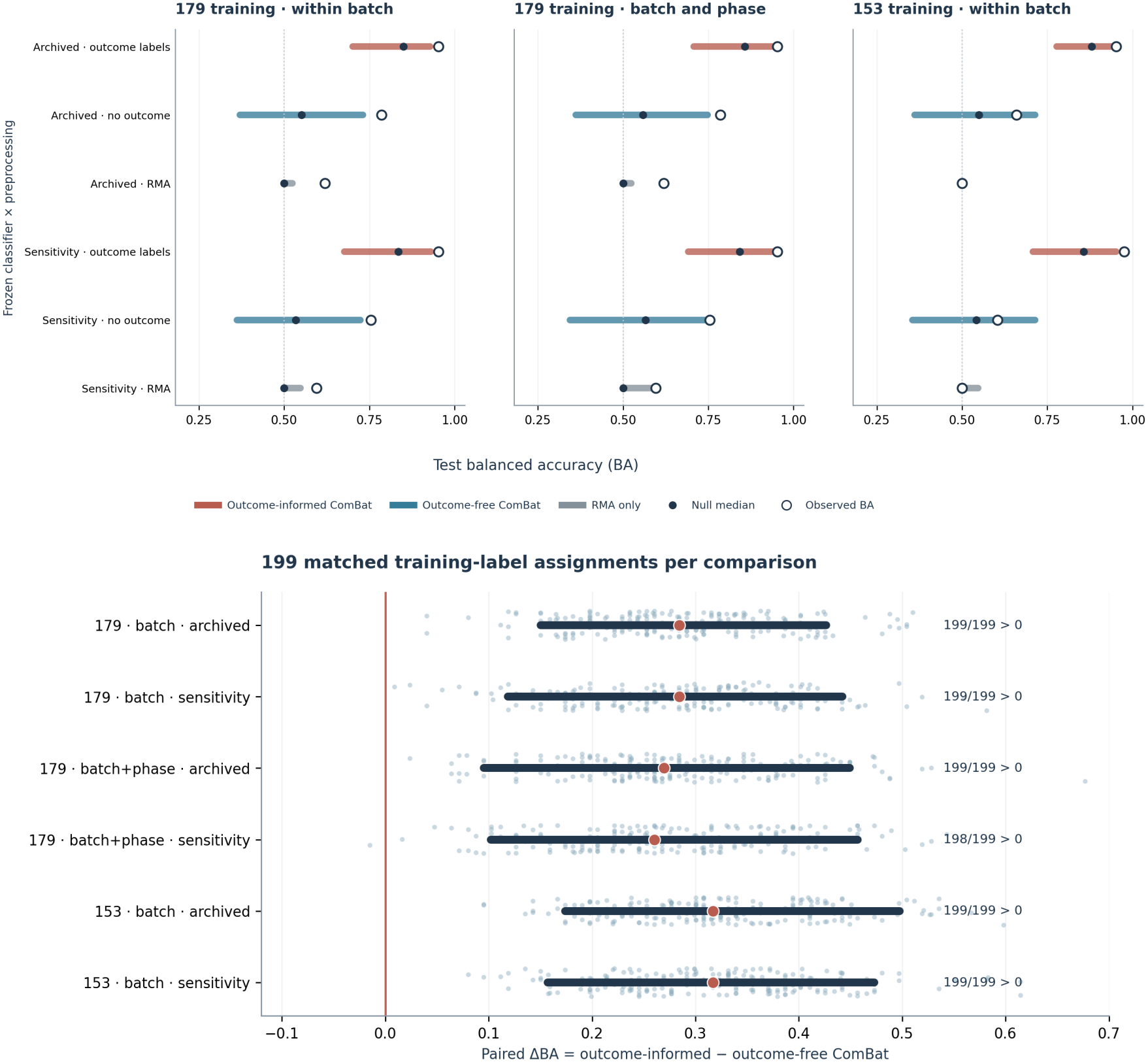
Training-label perturbations and apparent test balanced accuracy. Top: eighteen eligible 199-refit conditions under two fixed parameter sets, three processing strategies, and available stratification designs. Bottom: six paired outcome-informed-versus-outcome-free ComBat contrasts using exactly the same reassigned training labels. Percentile intervals summarize interventions on one reused test cohort; they are not future-patient uncertainty intervals.

The interpretation is deliberately limited. Preserving case totals within batches and phases can leave source-related information in the reassigned training data, and outcome-informed ComBat also changes which expression differences are preserved. The result establishes strong dependence of apparent performance on the information supplied during preprocessing. It does *not* prove that a future-patient classifier would have balanced accuracy 0.549, or that 0.881 minus 0.549 is a causal estimate of inflation in the original paper.

### 3.4 Published reuse is broader than one classifier, but the evidence has different grades

The targeted literature screen identified 29 records with search matches to both source series. Importantly, a shared mention is not evidence that the same measurements crossed a train/test split. The strongest publication-specific evidence outside the core reconstruction was a published stage-analysis sample list from Zhong et al. (2021): six GSE51981/GSE6364 pairs in the same listed group have identical CEL contents. This documents repeated analytical inclusion, which differs from mislabeling a train/test split; we did not quantify its effect on the published immune-infiltration analysis.

Several diagnostic publications placed GSE51981 and GSE6364 in opposite roles. Chen et al. (2023) report training with 76 GSE51981 endometriosis cases and evaluating GSE6364. If those 76 were selected from the same 77 GSE51981 ENDO arrays audited here, at most one of the 19 duplicated case arrays could have been left out, implying at least 18 potential cross-split copies. This is a *conditional count bound*, not a confirmed 18-GSM author training roster. Huang et al. (2022) report using GSE6364 plus GSE25628 to develop a signature and GSE51981 for evaluation, but the exact GSE51981 testing list has not been authenticated. Two further studies used these series on opposite sides of a reported training–evaluation split: Wei et al. (2023) trained with GSE51981 plus GSE25628 and evaluated with GSE6364 plus GSE7305 and GSE7307, whereas Yang et al. (2026) placed GSE6364 in a pooled training cohort and GSE51981 in validation. Both used additional datasets, and neither author-specific GSM partition has been independently authenticated here (Table 2). We avoid attributing intent or converting 29 indexed hits into a prevalence of leakage.

**Table 2.**
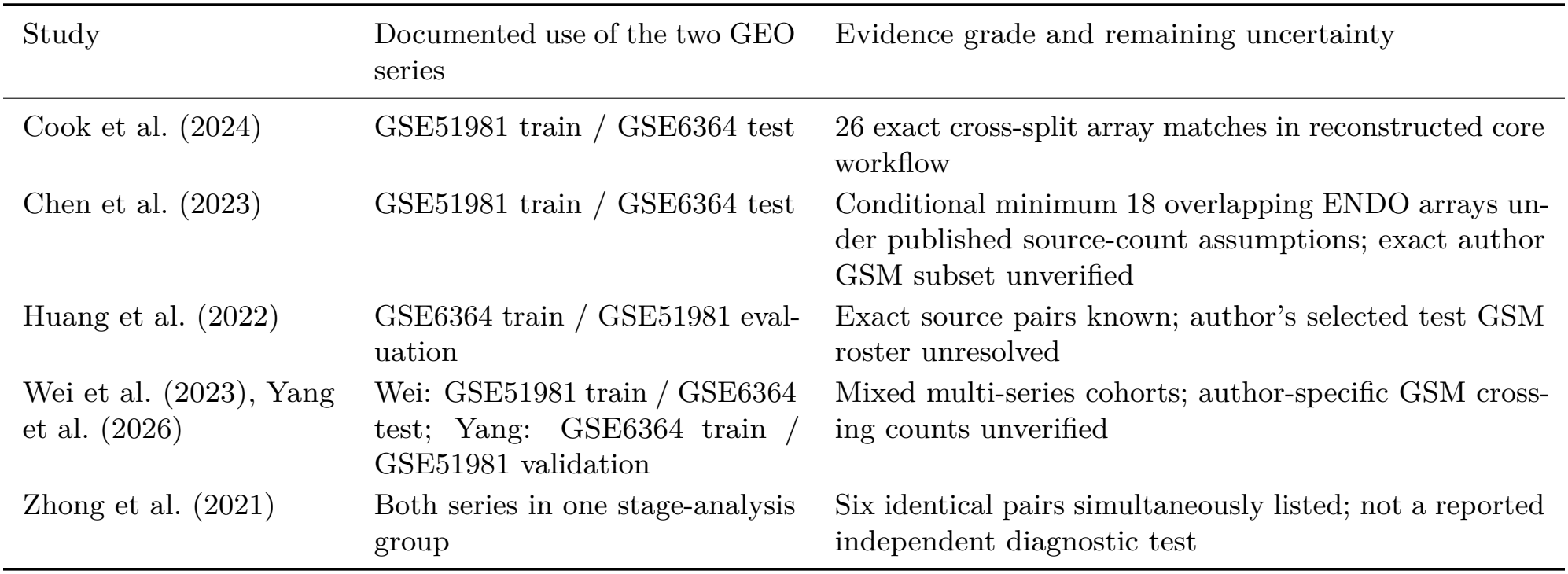
What can and cannot be established about reuse in the published record.

| Study | Documented use of the two GEO series | Evidence grade and remaining uncertainty |
|---|---|---|
| Cook et al. (2024) | GSE51981 train / GSE6364 test | 26 exact cross-split array matches in reconstructed core workflow |
| Chen et al. (2023) | GSE51981 train / GSE6364 test | Conditional minimum 18 overlapping ENDO arrays un-der published source-count assumptions; exact author GSM subset unverified |
| Huang et al. (2022) | GSE6364 train / GSE51981 eval-uation | Exact source pairs known; author's selected test GSM roster unresolved |
| Wei et al. (2023), Yang et al. (2026) | Wei: GSE51981 train / GSE6364 test; Yang: GSE6364 train / GSE51981 validation | Mixed multi-series cohorts; author-specific GSM cross-ing counts unverified |
| Zhong et al. (2021) | Both series in one stage-analysis group | Six identical pairs simultaneously listed; not a reported independent diagnostic test |

### 3.5 Study source and scanner date can strongly track disease labels

After training-side deduplication, the source-label table contained 56 cases and 62 controls from GSE51981, 20 controls only from GSE29981, and 15 controls only from GSE4888. Thus all retained training cases came from a single study. In a label-blind comparison of 324 fixed probe signals, 89.5% of probes had larger absolute differences between GSE29981 and GSE51981 controls than between cases and controls within GSE51981. The contrast reflects distinct tissue preparations and source effects; it does not establish that 89.5% of the classifier features were spurious.

GSE51981’s five scanner years also had markedly different clinical-group composition. In 2009, there were 46 ENDO and 31 OTHER arrays, but no NUPP controls. In 2010, there were only two ENDO arrays compared with 15 NUPP controls. The resulting calendar structure was sufficiently informative that an out-of-fold *scanner-date-only* control reached AUC **0.9347** for ENDO versus NUPP. The same samples’ 324-probe model reached AUC **0.8934** (Figure 4). The date control used only metadata from the training folds, so this finding is not due to scoring observations on the dates from which their labels were learned. It is a post-hoc technical negative control, not a proposed diagnostic predictor.

**Figure 4:**
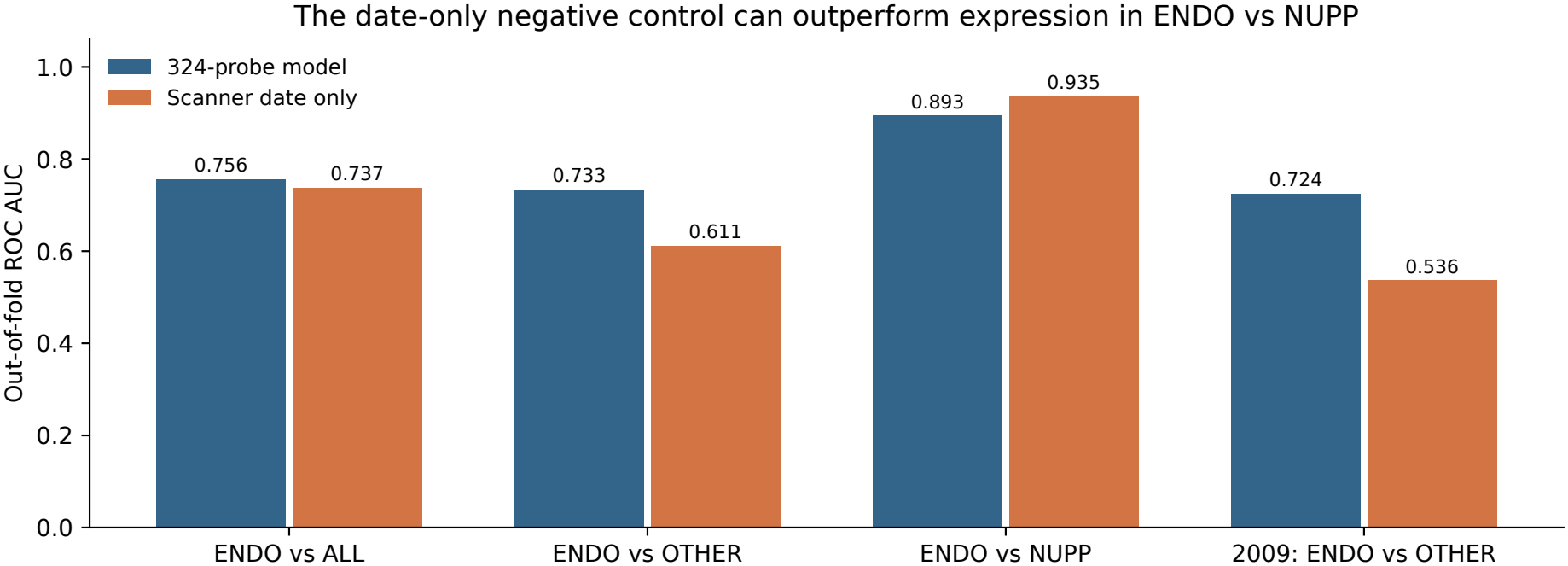
A technical negative control can outperform the molecular score. Out-of-fold date-only and molecular AUCs for the three diagnostic tasks and the restricted 2009 comparison. The dates of held-out arrays were seen only as covariates; their diagnoses were not used to construct the held-out date-only predictions. This analysis was added after the original benchmark.

A comparison of fixed molecular scores only among case–control pairs scanned on the same day gave rank concordance 0.677 for ENDO versus NUPP (62 eligible pairs). That value is neither the AUC of a retrained date-adjusted model nor evidence that the true date-free AUC equals 0.677. It shows that the apparently excellent full-sample separation cannot be read as independent evidence for a disease-specific gene signature without taking the calendar allocation seriously.

In a separate post-hoc analysis of ten previously published marker genes, apparent changes after date restriction were not always explained by statistical date adjustment. For example, the ENDO/NUPP log_2_ contrast for ACKR1 was +2.347 on all arrays, +1.121 when limited to shared acquisition dates, and +1.173 with date fixed effects fitted on that same restricted sample. For ENDO/OTHER, PDK3 changed from +0.206 on all arrays to *−* 0.186 on shared dates, remaining *−* 0.171 after date adjustment. Only LMNB1 in ENDO/NUPP met Holm correction across the 20 fixed-effect tests (adjusted *p* = 0.030). These results demonstrate the need to distinguish a change in comparison populations from a change in statistical adjustment; they do not invalidate the original multigene models (Supplementary Section S11).

### 3.6 A more contained internal benchmark retains moderate but limited discrimination

The new within-GSE51981 benchmark had out-of-fold AUC 0.7556 for ENDO versus all non-ENDO, 0.7329 for ENDO versus OTHER, and 0.8934 for ENDO versus NUPP (Table 3). Conditional sample-bootstrap intervals were 0.671–0.836, 0.625–0.829 and 0.823–0.952, respectively. The ENDO-versus-OTHER question is arguably more relevant to differential diagnosis than a comparison with only strict controls, but the model’s balanced accuracy there was 0.643 and its Brier score (0.2198) did not improve on the phase-only baseline (0.2168). Ranking performance should not be mistaken for calibrated clinical risk probabilities.

**Table 3.** Exploratory sample-level internal performance of an annotation-derived 324-probe classifier.

| Task (ENDO/controls) | ROC AUC (95% CI) | Balanced accuracy | Brier |
|---|---|---|---|
| All non-ENDO (77/71) | 0.756 (0.671–0.836) | 0.702 | 0.2200 |
| OTHER (77/37) | 0.733 (0.625–0.829) | 0.643 | 0.2198 |
| NUPP (77/34) | 0.893 (0.823–0.952) | 0.803 | 0.1459 |
| OTHER, 2009 only (46/31) | 0.724 (0.605–0.836) | 0.622 | 0.2611 |

Out-of-fold predictions from a newly trained model, not the original Fogg classifier. Confidence intervals resample fixed sample-level scores. Donor correspondence and the historical feature-rule selection process remain incomplete.

Restricting evaluation to 2009, when both ENDO and OTHER were represented, gave AUC 0.7237 (46/31). A post-hoc leave-one-scanner-date-out analysis within the same year gave pooled AUC 0.6978 and a mean of the five held-out-date AUCs of 0.6775. The corresponding empirical add-one fractions under within-date training-label permutations were 0.030 and 0.070; these are conditional permutation results, not confirmation of prospective patient-level validity. The two different date summaries weight test dates differently, and several dates contain few controls. We therefore report both rather than selecting the more favorable one.

### 3.7 Exact-copy exposure changes the five-gene RF reconstruction

The Huang-inspired forest scored the 26 duplicate GSE51981 arrays perfectly by rank when their identical CEL contents had been included in the 59-array training set (AUC 1.000). On the other 85 ENDO/NUPP arrays without exact training copies, AUC was 0.8094. This raw difference could reflect many properties of the samples, including scanner date, so we did not interpret it as the effect of duplication by itself.

**Figure 5:**
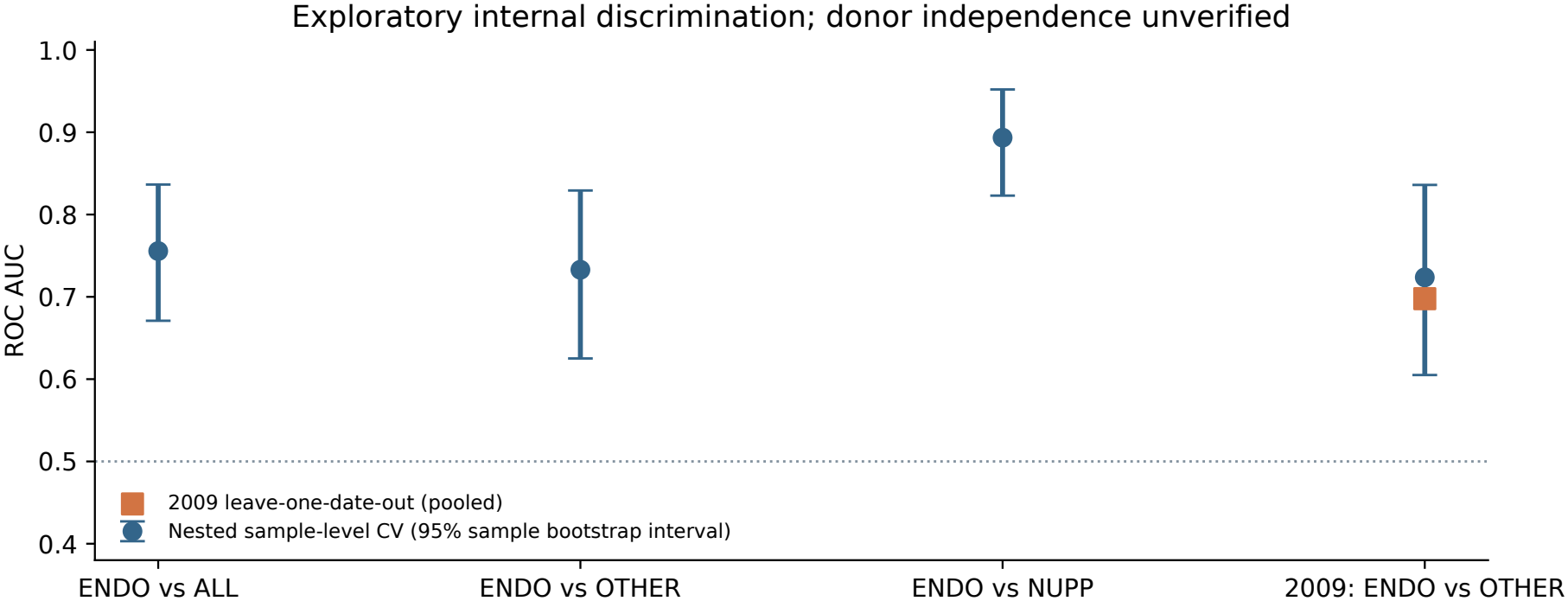
Internal molecular evaluation under more contained fitting. Out-of-fold AUC and fixed-score confidence intervals for three control definitions and a 2009-only comparison. The separately reported leave-one-date-out statistic addresses a different validation question and is not an external patient-level test.

**Figure 6:**
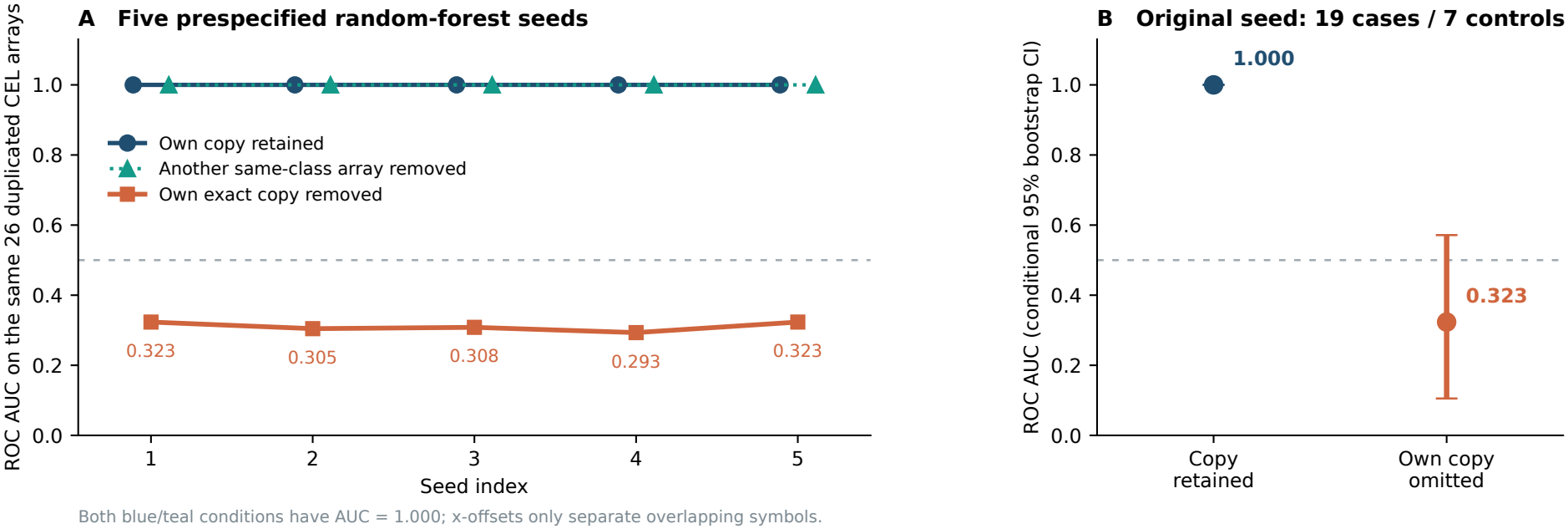
Same test arrays, different training exposure. A five-gene Huang-inspired random-forest reconstruction scored 26 repeated CEL arrays with or without the target’s own exact training copy. In panel A, the “copy present” and “other same-class array removed” curves coincide at AUC 1.000 across all five seeds. Each own-copy-out score came from a separate 58-sample fit; these are intervention sensitivities, not estimates for the original author-trained model.

Instead we held every test array fixed and refitted the same 1,000-tree random-forest specification after removing *only its own* exact training counterpart. The 26 resulting target-specific predictions yielded AUC **0.3233** (conditional 95% stratified sample-bootstrap interval **0.1053–0.5714**). With a different, same-class GSE6364 training array removed, retaining the target’s copy, the corresponding aggregated control score AUC remained 1.000. Thus the training-set size was 58 in both comparisons; the crucial difference was whether the exact target measurement remained exposed to fitting.

The own-copy-out result was similar across five predeclared forest seeds (AUC 0.2932–0.3233), while all five exact-copy-exposed evaluations retained AUC 1.000. Because the interval for 0.3233 includes 0.5, we do not claim statistically established below-chance performance. Predictions were pooled from separately fitted forests and are not an independent clinical test of one new model. Across all 111 ENDO/NUPP records, a *hybrid* score set in which only duplicate targets received own-copy-out predictions had AUC 0.7086 versus 0.8657 in the original reconstruction; across all 148 ENDO/non-ENDO records, the corresponding values were 0.6116 versus 0.7545. These hybrid comparisons are illustrative, not numerical corrections to Huang et al. (2022)’s published AUC of 0.656.

## 4 Discussion

### 4.1 What the audit adds beyond documenting an identical file

The central problem is not that public datasets have been reused. The same CEL file can be valuable for biological replication of a method, technical quality control, or reinterpretation under a new question. The problem arises when its reappearance is missed and the file counts as evidence from an independent test patient. By comparing decompressed raw arrays, we established 26 exact cross-series identities without setting an arbitrary expression-similarity threshold. The clustering of those identities in 2004–2005 also helps explain why source reuse and calendar effects cannot be discussed as unrelated properties of this particular dataset.

The ComBat analyses address a different information path. The conditional diagnosis of a nominal test sample was passed into a joint preprocessing model before that sample was scored. The one-label interventions show the resulting features and predictions are responsive to that supplied diagnosis. More unusually, the large post-hoc re-training experiment shows that high nominal performance can survive extensive rearrangement of individual training diagnoses so long as the true test diagnoses still enter ComBat. This pattern is compelling evidence of test-target dependence. It is not a controlled experiment that decomposes the numerical accuracy of the authors’ original fitted classifier into a biological component plus a leakage component.

The five-gene reconstruction supplies a complementary intervention: a target’s exact training array can be removed while leaving the same target and almost the entire training cohort in place. The strong change in pooled rank discrimination and its stability across seeds make the training-copy dependence difficult to dismiss as an accidental feature of one forest seed. At the same time, the reconstruction used different signal processing from the original Huang study, and each own-copy-out prediction came from a different fit. We therefore resist the tempting but unsupported statement that a published external AUC has been *corrected* by 0.677.

### 4.2 Calendar composition can materially affect molecular classification

The high date-only AUC illustrates how powerfully a technical acquisition characteristic can track diagnosis in a retrospectively assembled cohort. For ENDO versus NUPP, the 2009 period contained many cases but no strict controls, whereas 2010 contained mainly controls. This is a problem of study design and sampling support, not proof that date itself causes disease or that no endometriosis biology was measured. Within GSE51981, the more clinically relevant ENDO-versus-OTHER comparison, particularly in 2009, retained moderate molecular discrimination, but it also had small effective sample counts per acquisition date and limited probability calibration.

### 4.3 A practical response for investigators reusing GEO data

Prediction-model reporting and risk-of-bias guidance emphasizes clear descriptions of study design, model development, evaluation, and applicability (Collins et al., 2024; Moons et al., 2025). A credible validation study needs more than a train/test column. It needs a specimen- and file-level provenance map, a clear statement of which labels were available at each stage of processing, and a split that cannot place a known repeated measurement on both sides. It should also distinguish performance within one acquisition setting from performance in a new cohort with a genuinely unseen batch. A transductive correction that has no target diagnoses is different from an inductive algorithm that works on a single new patient’s measurement.

We have organized these checks into a proposed seven-item *GEO-LEAK* reporting checklist and a small disease-agnostic research-preview utility, ArrayProvenance. The research preview can report exact CEL hash clusters, known split conflicts, source/date composition and unresolved donor fields. It is currently a research preview, rather than a validated diagnostic tool. Its contribution is practical transparency: the output should explicitly say *unknown* when a donor link or a public author roster cannot be verified, rather than interpreting a different GSM accession as an independent patient.

## 5 Limitations and scope of claims

Several constraints remain. The donor-level crosswalk for all 148 GSE51981 arrays was not authenticated, so the internal cross-validation is sample-level and not a certified independent-patient analysis. The 193-gene panel can be rederived from annotation without using diagnoses, but that reconstruction alone does not document the complete historical timeline of feature-rule choice. The fixed out-of-fold bootstrap intervals do not cover all model-training or selection uncertainty. The date-only controls and leave-one-date-out analysis were added after the primary internal benchmark and are interpreted as post-hoc sensitivity checks.

The original fitted Huang forest was unavailable, and our five-gene RF is not its exact reproduction; similarly, matching the Fogg core confusion matrix does not establish identity of every original author coefficient or score. In the published-reuse registry, the precise GSM partition for several papers remains unknown; the conditional Chen lower bound and the confirmed simultaneous Zhong duplicates describe different evidential situations. The 29 dual-hit papers are not a representative sample of all endometriosis classifiers. Source, date, tissue and disease may be confounded, and no sensitivity analysis here uniquely identifies their causal contributions. We did not establish discrimination in an independent new clinical cohort, test deployment on unseen batches, or demonstrate clinical benefit.

Finally, the important biological observations of the original studies are not negated by finding a weakness in their diagnostic evaluation design. An expression association can remain scientifically valuable even when a claimed external discrimination statistic is not interpretable as independent validation. Our conclusions concern specific pipelines and the assumptions used to assess them, not the integrity of the researchers who first generated or reused the public arrays.

## 5 Conclusion

Across widely reused endometriosis microarray series, nominally different GEO accessions masked 26 identical CEL measurements. In a reconstructed matrisome workflow, true held-out diagnoses entered ComBat and could influence the downstream classifier, including after training-array deduplication and training-label rearrangement. In a separately reconstructed five-gene forest, removing only the target’s own exact training copy caused a large, reproducible shift in ranked predictions on unchanged test arrays. A more contained internal evaluation retained moderate discrimination against other gynecological conditions but remained sensitive to how controls and acquisition dates were defined. The practical lesson is simple: independent diagnosis cannot be established from accession numbers or high AUC alone. Source identity, target information flow, and the intended setting for future predictions must all be audited.

## Data Availability

All transcriptomic data analyzed in this study were obtained from publicly available repositories, including the NCBI Gene Expression Omnibus (GEO). Relevant dataset accession numbers and analysis details are provided in the manuscript and Supplementary Material. Derived results and supplementary analyses are included in the accompanying supplementary files. No new human samples or participant-level clinical data were collected for this study.

https://www.ncbi.nlm.nih.gov/geo/query/acc.cgi?acc=GSE51981

https://www.ncbi.nlm.nih.gov/geo/query/acc.cgi?acc=GSE6364

## Data and code availability

Raw CEL measurements were obtained from the publicly accessible GEO series GSE4888, GSE29981, GSE51981, GSE6364 and GSE25628; no raw CEL archive is redistributed. The accompanying supplementary data contain the source hash manifest, 26-pair CEL identity crosswalk, reconstructed sample partitions, literature screen, and intervention and benchmark tables. Supplementary code contains the analytical scripts and figure-generation sources used for this manuscript. The Fogg Lab’s original publicly released code is cited separately (Fogg Lab, 2023). ArrayProvenance is a research-preview implementation of the proposed provenance checks; no public DOI or software release is claimed in this version.

## Ethics and declarations

### Ethics

This secondary computational analysis used publicly available gene-expression data and metadata. No new participants were recruited and no new specimens were collected.

### Author contributions

Elena Krikun performed the computational investigation, verified the archived outputs, interpreted the results, and drafted the original manuscript. Abedalrhman Alkhateeb reviewed and edited the manuscript.

## Acknowledgments

We thank the investigators who generated and openly shared the source transcriptomic datasets and analysis code.

## Use of generative AI and AI-assisted technologies

OpenAI ChatGPT and Anthropic Claude were used to assist with analysis planning, and language editing. The authors are responsible for reviewing the computational outputs and scientific interpretations and accept responsibility for the reported methods, claims, and manuscript. No AI system is listed as an author.

