## supplemental pdf for "Hidden Dependencies in Endometriosis Microarray Classifiers: Sample Reuse, Outcome-Informed Batch Correction, and Diagnostic Reassessment"

### Supplementary Information
### Hidden Dependencies in Endometriosis Microarray Classifiers

Elena Krikun[1,*] Abedalrhman Alkhateeb[2]

[1]Lakehead University, Thunder Bay, Ontario, Canada

[2]Department of Computer Science, Lakehead University, Thunder Bay, Ontario, Canada

**Scope and evidence grades.** These tables preserve the complete earlier ComBat intervention audit (Tables S1–S6), including 1,194 paired refit comparisons in full-precision CSV, without replacing or silently changing those analyses. Later sections document the array/source/date audit, published reuse, the new internal benchmark, the reconstructed Huang sensitivity study, and the limits of each evidence source. Confirmed identity means a matching decompressed CEL SHA-256; it does not establish donor independence. Conditional inferences about article-specific sample rosters are labeled explicitly.

#### Contents

**Supplementary Table S1** 2

**Supplementary Table S2** 3

**Supplementary Table S3** 7

**Supplementary Table S4** 8

**Supplementary Table S5** 9

**Supplementary Table S6** 9

**S7. Source, phase, and scanner support** 10

**S8. Internal model and date controls** 10

**S9. Held-out-date evaluation** 10

**S10. Huang reconstruction** 11

**S11. Marker panel and scan dates** 11

**S12. Literature and GEO-LEAK** 13

**S13. Supplementary graphics** 13

### Supplementary Table S1. Byte-identical CEL arrays found in training and test

| Test GSM | Train GSM | Case | Uncompressed CEL SHA-256 | Bytes |
|---|---|---|---|---|
| GSM150190 | GSM1256666 | 1 | 5d61b91977b50a5b3a8cf7ba85cb83c746d68c9824c14d1b0751db086d4e4ede | 13551381 |
| GSM150191 | GSM1256655 | 1 | 561b8ed69ca9cf13414996d9caaf06f008913d0893651d20bb9d53187546d9bb | 13551336 |
| GSM150193 | GSM1256660 | 1 | b47ac97ee858b0cbbf0fff1be91b6ef8719de02a2b3a7c48c30a7dd9e02c5c12 | 13550849 |
| GSM150194 | GSM1256663 | 1 | eab961fc5a593a8a4fbd2d10190be3c8231d1a94420bce99fabc80ce33e08050 | 13550817 |
| GSM150195 | GSM1256664 | 1 | b835f9bbef3ecbbe0ba27e4b8f2cff71f94964dae1d0552cc641c4964a7f3f27 | 13551137 |
| GSM150198 | GSM1256770 | 0 | 9ff7502e58ba0b240120ffc8633f1aab77622a282ae719bbf026b5d31f086147 | 13551126 |
| GSM150199 | GSM1256771 | 0 | 7bca93e6c1e6471615fe0c901d7005405312344999b0bde41f2fed7f4db76537 | 13552722 |
| GSM150201 | GSM1256772 | 0 | 845c85b931feba2388d02b6aaf5ffff9a9264661c2faa30900bc875c225a4d9c | 13551126 |
| GSM150202 | GSM1256667 | 1 | 52d99c9011625e5cc1ab33147dfe98a3a2d88dc20a63e3b3c32c436c8ee7f2f4 | 13551905 |
| GSM150203 | GSM1256653 | 1 | 50eee14c217c7c6ff9031a61c9ea7d7b94462a6722af4917e38740dfaf0d09d5 | 13551365 |
| GSM150204 | GSM1256654 | 1 | d7771864d58e57b4054c3fd471a63e7dac3f06042fc022dcecfeb0a889de84b3 | 13550909 |
| GSM150205 | GSM1256657 | 1 | b4a64bdbfee3039073ab38ffbbbb5846291f75a3a9a9afa6ee89ca2cf954fd0b | 13550700 |
| GSM150207 | GSM1256661 | 1 | 71e12b5786f766a383eb6ac3fbdbe0206335230cf860956794b4e8f0b387ebd5 | 13551817 |
| GSM150211 | GSM1256668 | 1 | df789ff8ef995f7c64dbe916e84812a1ac3efb193757a83403469d926a0532e7 | 13550872 |
| GSM150212 | GSM1256656 | 1 | 674f43300c773ff5efcf5e28f17f9c146b79135a7b8d88460134c307eecf7108 | 13550837 |
| GSM150213 | GSM1256658 | 1 | f4b2e8835bebd107a078abe8e75006c20f9b2c2091cf830f480c9b4219f2c811 | 13550285 |
| GSM150214 | GSM1256659 | 1 | 5715168977d41b71bcb7aba15981796b0bd5950971767a1015fa495b17defee7 | 13551656 |
| GSM150215 | GSM1256662 | 1 | 1f035e9bb3305a5f2dfdd11f0a263be5ce9466f18b6e30c8bd5ca8b4b51fde82 | 13550753 |
| GSM150216 | GSM1256665 | 1 | 8ae5e38beb90751f26d7d75664214ff54c7264e6aaadc8d2e318d4966f3881bc | 13551805 |
| GSM150217 | GSM1256669 | 1 | 47177fac72731b59598869b9cd376cecbf08d5f8ac2a88357a01a4e252e0ef86 | 13551337 |
| GSM150218 | GSM1256670 | 1 | 72f0a68d318dc261743b3835a7769b0d7a3048be52251c87c89145619f58b739 | 13551146 |
| GSM150219 | GSM1256671 | 1 | 91838c51600941d9120334c348615f3f556ef5b107e24f7f5569e7108c19c95c | 13550922 |
| GSM150224 | GSM1256766 | 0 | eda50c4b6801ccd56c7ca0c656e383aec0cc846aec156d5a21e9f9e097e51436 | 13551838 |
| GSM150225 | GSM1256767 | 0 | 53ba12f07897286fa59fc16535770591d39e181fb111cf8b0fe73e2f53e474de | 13550826 |
| GSM150226 | GSM1256768 | 0 | a2f386210ddb8f1434a254f18d08f764c35db8548952580d983626f085347118 | 13551114 |
| GSM150227 | GSM1256769 | 0 | c035549e6afff8bed948ac83da637bedfdc29db7d87fae17f022e774f4835115 | 13550890 |

*All 26 pairs were verified using SHA-256 hashes of decompressed CEL files. The accompanying CSV also provides the full archive-member names.*

#### Supplementary Table S2. The 216 arrays used in the reconstructed classification workflow

| GSM | Batch | Case | Partition | Menstrual phase |
|---|---|---|---|---|
| GSM109814 | 1 | 0 | TRAIN179 | proliferative |
| GSM109815 | 1 | 0 | TRAIN179 | proliferative |
| GSM109816 | 1 | 0 | TRAIN179 | proliferative |
| GSM109817 | 1 | 0 | TRAIN179 | proliferative |
| GSM109820 | 1 | 0 | TRAIN179 | early_secretory |
| GSM109821 | 1 | 0 | TRAIN179 | early_secretory |
| GSM109822 | 1 | 0 | TRAIN179 | early_secretory |
| GSM109824 | 1 | 0 | TRAIN179 | mid_secretory |
| GSM109825 | 1 | 0 | TRAIN179 | mid_secretory |
| GSM109826 | 1 | 0 | TRAIN179 | mid_secretory |
| GSM109827 | 1 | 0 | TRAIN179 | mid_secretory |
| GSM109828 | 1 | 0 | TRAIN179 | mid_secretory |
| GSM109829 | 1 | 0 | TRAIN179 | mid_secretory |
| GSM109830 | 1 | 0 | TRAIN179 | mid_secretory |
| GSM109831 | 1 | 0 | TRAIN179 | mid_secretory |
| GSM1256653 | 3 | 1 | TRAIN179 | early_secretory |
| GSM1256654 | 3 | 1 | TRAIN179 | early_secretory |
| GSM1256655 | 3 | 1 | TRAIN179 | proliferative |
| GSM1256656 | 3 | 1 | TRAIN179 | mid_secretory |
| GSM1256657 | 3 | 1 | TRAIN179 | early_secretory |
| GSM1256658 | 3 | 1 | TRAIN179 | mid_secretory |
| GSM1256659 | 3 | 1 | TRAIN179 | mid_secretory |
| GSM1256660 | 3 | 1 | TRAIN179 | proliferative |
| GSM1256661 | 3 | 1 | TRAIN179 | early_secretory |
| GSM1256662 | 3 | 1 | TRAIN179 | mid_secretory |
| GSM1256663 | 3 | 1 | TRAIN179 | proliferative |
| GSM1256664 | 3 | 1 | TRAIN179 | proliferative |
| GSM1256665 | 3 | 1 | TRAIN179 | mid_secretory |
| GSM1256666 | 3 | 1 | TRAIN179 | proliferative |
| GSM1256667 | 3 | 1 | TRAIN179 | early_secretory |
| GSM1256668 | 3 | 1 | TRAIN179 | mid_secretory |
| GSM1256669 | 3 | 1 | TRAIN179 | mid_secretory |
| GSM1256670 | 3 | 1 | TRAIN179 | mid_secretory |
| GSM1256671 | 3 | 1 | TRAIN179 | mid_secretory |
| GSM1256672 | 3 | 1 | TRAIN179 | mid_secretory |
| GSM1256673 | 3 | 1 | TRAIN179 | proliferative |
| GSM1256674 | 3 | 1 | TRAIN179 | mid_secretory |
| GSM1256675 | 3 | 1 | TRAIN179 | mid_secretory |
| GSM1256676 | 3 | 1 | TRAIN179 | mid_secretory |
| GSM1256677 | 3 | 1 | TRAIN179 | mid_secretory |
| GSM1256678 | 3 | 1 | TRAIN179 | proliferative |
| GSM1256679 | 3 | 1 | TRAIN179 | proliferative |
| GSM1256680 | 3 | 1 | TRAIN179 | proliferative |
| GSM1256681 | 3 | 1 | TRAIN179 | early_secretory |
| GSM1256682 | 3 | 1 | TRAIN179 | proliferative |
| GSM1256683 | 3 | 1 | TRAIN179 | proliferative |
| GSM1256684 | 3 | 1 | TRAIN179 | proliferative |
| GSM1256685 | 3 | 1 | TRAIN179 | proliferative |
| GSM1256686 | 3 | 1 | TRAIN179 | proliferative |
| GSM1256687 | 3 | 1 | TRAIN179 | early_secretory |
| GSM1256688 | 3 | 1 | TRAIN179 | mid_secretory |
| GSM1256689 | 3 | 1 | TRAIN179 | proliferative |

Table S2 (continued)

| GSM | Batch | Case | Partition | Menstrual phase |
|---|---|---|---|---|
| GSM1256690 | 3 | 1 | TRAIN179 | mid_secretory |
| GSM1256691 | 3 | 1 | TRAIN179 | proliferative |
| GSM1256692 | 3 | 1 | TRAIN179 | mid_secretory |
| GSM1256693 | 3 | 1 | TRAIN179 | proliferative |
| GSM1256694 | 3 | 1 | TRAIN179 | mid_secretory |
| GSM1256695 | 3 | 1 | TRAIN179 | early_secretory |
| GSM1256696 | 3 | 1 | TRAIN179 | mid_secretory |
| GSM1256697 | 3 | 1 | TRAIN179 | mid_secretory |
| GSM1256698 | 3 | 1 | TRAIN179 | proliferative |
| GSM1256699 | 3 | 1 | TRAIN179 | proliferative |
| GSM1256700 | 3 | 1 | TRAIN179 | mid_secretory |
| GSM1256701 | 3 | 1 | TRAIN179 | early_secretory |
| GSM1256702 | 3 | 1 | TRAIN179 | proliferative |
| GSM1256703 | 3 | 1 | TRAIN179 | proliferative |
| GSM1256704 | 3 | 1 | TRAIN179 | mid_secretory |
| GSM1256705 | 3 | 1 | TRAIN179 | early_secretory |
| GSM1256706 | 3 | 1 | TRAIN179 | proliferative |
| GSM1256707 | 3 | 1 | TRAIN179 | proliferative |
| GSM1256708 | 3 | 1 | TRAIN179 | early_secretory |
| GSM1256709 | 3 | 1 | TRAIN179 | proliferative |
| GSM1256710 | 3 | 1 | TRAIN179 | mid_secretory |
| GSM1256711 | 3 | 1 | TRAIN179 | mid_secretory |
| GSM1256712 | 3 | 1 | TRAIN179 | early_secretory |
| GSM1256713 | 3 | 1 | TRAIN179 | early_secretory |
| GSM1256714 | 3 | 1 | TRAIN179 | mid_secretory |
| GSM1256715 | 3 | 1 | TRAIN179 | mid_secretory |
| GSM1256716 | 3 | 1 | TRAIN179 | early_secretory |
| GSM1256717 | 3 | 1 | TRAIN179 | early_secretory |
| GSM1256718 | 3 | 1 | TRAIN179 | proliferative |
| GSM1256719 | 3 | 1 | TRAIN179 | proliferative |
| GSM1256720 | 3 | 0 | TRAIN179 | proliferative |
| GSM1256721 | 3 | 0 | TRAIN179 | early_secretory |
| GSM1256722 | 3 | 0 | TRAIN179 | proliferative |
| GSM1256723 | 3 | 0 | TRAIN179 | proliferative |
| GSM1256724 | 3 | 0 | TRAIN179 | early_secretory |
| GSM1256725 | 3 | 0 | TRAIN179 | early_secretory |
| GSM1256726 | 3 | 0 | TRAIN179 | proliferative |
| GSM1256727 | 3 | 0 | TRAIN179 | early_secretory |
| GSM1256728 | 3 | 0 | TRAIN179 | mid_secretory |
| GSM1256729 | 3 | 0 | TRAIN179 | proliferative |
| GSM1256730 | 3 | 0 | TRAIN179 | early_secretory |
| GSM1256731 | 3 | 0 | TRAIN179 | proliferative |
| GSM1256732 | 3 | 0 | TRAIN179 | mid_secretory |
| GSM1256733 | 3 | 0 | TRAIN179 | proliferative |
| GSM1256734 | 3 | 0 | TRAIN179 | proliferative |
| GSM1256735 | 3 | 0 | TRAIN179 | mid_secretory |
| GSM1256736 | 3 | 0 | TRAIN179 | mid_secretory |
| GSM1256737 | 3 | 0 | TRAIN179 | mid_secretory |
| GSM1256738 | 3 | 0 | TRAIN179 | mid_secretory |
| GSM1256739 | 3 | 0 | TRAIN179 | proliferative |
| GSM1256740 | 3 | 0 | TRAIN179 | mid_secretory |
| GSM1256741 | 3 | 0 | TRAIN179 | proliferative |
| GSM1256742 | 3 | 0 | TRAIN179 | proliferative |
| GSM1256743 | 3 | 0 | TRAIN179 | mid_secretory |
| GSM1256744 | 3 | 0 | TRAIN179 | proliferative |
| GSM1256745 | 3 | 0 | TRAIN179 | proliferative |
| GSM1256746 | 3 | 0 | TRAIN179 | mid_secretory |
| GSM1256747 | 3 | 0 | TRAIN179 | mid_secretory |
| GSM1256748 | 3 | 0 | TRAIN179 | proliferative |
| GSM1256749 | 3 | 0 | TRAIN179 | mid_secretory |
| GSM1256750 | 3 | 0 | TRAIN179 | mid_secretory |

Table S2 (continued)

| GSM | Batch | Case | Partition | Menstrual phase |
|---|---|---|---|---|
| GSM1256751 | 3 | 0 | TRAIN179 | early_secretory |
| GSM1256752 | 3 | 0 | TRAIN179 | early_secretory |
| GSM1256753 | 3 | 0 | TRAIN179 | proliferative |
| GSM1256754 | 3 | 0 | TRAIN179 | proliferative |
| GSM1256755 | 3 | 0 | TRAIN179 | mid_secretory |
| GSM1256756 | 3 | 0 | TRAIN179 | proliferative |
| GSM1256757 | 3 | 0 | TRAIN179 | mid_secretory |
| GSM1256758 | 3 | 0 | TRAIN179 | mid_secretory |
| GSM1256759 | 3 | 0 | TRAIN179 | proliferative |
| GSM1256760 | 3 | 0 | TRAIN179 | proliferative |
| GSM1256761 | 3 | 0 | TRAIN179 | proliferative |
| GSM1256762 | 3 | 0 | TRAIN179 | proliferative |
| GSM1256763 | 3 | 0 | TRAIN179 | proliferative |
| GSM1256764 | 3 | 0 | TRAIN179 | early_secretory |
| GSM1256765 | 3 | 0 | TRAIN179 | early_secretory |
| GSM1256766 | 3 | 0 | TRAIN179 | mid_secretory |
| GSM1256767 | 3 | 0 | TRAIN179 | mid_secretory |
| GSM1256768 | 3 | 0 | TRAIN179 | mid_secretory |
| GSM1256769 | 3 | 0 | TRAIN179 | mid_secretory |
| GSM1256770 | 3 | 0 | TRAIN179 | proliferative |
| GSM1256771 | 3 | 0 | TRAIN179 | proliferative |
| GSM1256772 | 3 | 0 | TRAIN179 | proliferative |
| GSM1256774 | 3 | 1 | TRAIN179 | early_secretory |
| GSM1256776 | 3 | 1 | TRAIN179 | early_secretory |
| GSM1256777 | 3 | 1 | TRAIN179 | proliferative |
| GSM1256778 | 3 | 1 | TRAIN179 | early_secretory |
| GSM1256779 | 3 | 1 | TRAIN179 | proliferative |
| GSM1256780 | 3 | 1 | TRAIN179 | mid_secretory |
| GSM1256781 | 3 | 1 | TRAIN179 | mid_secretory |
| GSM1256782 | 3 | 1 | TRAIN179 | proliferative |
| GSM1256784 | 3 | 0 | TRAIN179 | proliferative |
| GSM1256785 | 3 | 0 | TRAIN179 | proliferative |
| GSM1256786 | 3 | 0 | TRAIN179 | proliferative |
| GSM1256787 | 3 | 0 | TRAIN179 | early_secretory |
| GSM1256788 | 3 | 0 | TRAIN179 | proliferative |
| GSM1256789 | 3 | 0 | TRAIN179 | mid_secretory |
| GSM1256790 | 3 | 0 | TRAIN179 | early_secretory |
| GSM1256791 | 3 | 0 | TRAIN179 | mid_secretory |
| GSM1256792 | 3 | 0 | TRAIN179 | early_secretory |
| GSM1256793 | 3 | 0 | TRAIN179 | proliferative |
| GSM1256794 | 3 | 0 | TRAIN179 | proliferative |
| GSM1256796 | 3 | 0 | TRAIN179 | proliferative |
| GSM1256797 | 3 | 0 | TRAIN179 | mid_secretory |
| GSM1256798 | 3 | 0 | TRAIN179 | proliferative |
| GSM1256799 | 3 | 0 | TRAIN179 | proliferative |
| GSM1256800 | 3 | 0 | TRAIN179 | proliferative |
| GSM150190 | 2 | 1 | TEST37 | |
| GSM150191 | 2 | 1 | TEST37 | |
| GSM150192 | 2 | 1 | TEST37 | |
| GSM150193 | 2 | 1 | TEST37 | |
| GSM150194 | 2 | 1 | TEST37 | |
| GSM150195 | 2 | 1 | TEST37 | |
| GSM150196 | 2 | 0 | TEST37 | |
| GSM150197 | 2 | 0 | TEST37 | |
| GSM150198 | 2 | 0 | TEST37 | |
| GSM150199 | 2 | 0 | TEST37 | |
| GSM150201 | 2 | 0 | TEST37 | |
| GSM150202 | 2 | 1 | TEST37 | |
| GSM150203 | 2 | 1 | TEST37 | |
| GSM150204 | 2 | 1 | TEST37 | |
| GSM150205 | 2 | 1 | TEST37 | |

Table S2 (continued)

| GSM | Batch | Case | Partition | Menstrual phase |
|---|---|---|---|---|
| GSM150206 | 2 | 1 | TEST37 | |
| GSM150207 | 2 | 1 | TEST37 | |
| GSM150208 | 2 | 0 | TEST37 | |
| GSM150209 | 2 | 0 | TEST37 | |
| GSM150210 | 2 | 0 | TEST37 | |
| GSM150211 | 2 | 1 | TEST37 | |
| GSM150212 | 2 | 1 | TEST37 | |
| GSM150213 | 2 | 1 | TEST37 | |
| GSM150214 | 2 | 1 | TEST37 | |
| GSM150215 | 2 | 1 | TEST37 | |
| GSM150216 | 2 | 1 | TEST37 | |
| GSM150217 | 2 | 1 | TEST37 | |
| GSM150218 | 2 | 1 | TEST37 | |
| GSM150219 | 2 | 1 | TEST37 | |
| GSM150220 | 2 | 0 | TEST37 | |
| GSM150221 | 2 | 0 | TEST37 | |
| GSM150222 | 2 | 0 | TEST37 | |
| GSM150223 | 2 | 0 | TEST37 | |
| GSM150224 | 2 | 0 | TEST37 | |
| GSM150225 | 2 | 0 | TEST37 | |
| GSM150226 | 2 | 0 | TEST37 | |
| GSM150227 | 2 | 0 | TEST37 | |
| GSM742055 | 4 | 0 | TRAIN179 | proliferative |
| GSM742057 | 4 | 0 | TRAIN179 | proliferative |
| GSM742058 | 4 | 0 | TRAIN179 | proliferative |
| GSM742061 | 4 | 0 | TRAIN179 | early_secretory |
| GSM742063 | 4 | 0 | TRAIN179 | mid_secretory |
| GSM742065 | 4 | 0 | TRAIN179 | proliferative |
| GSM742067 | 4 | 0 | TRAIN179 | early_secretory |
| GSM742068 | 4 | 0 | TRAIN179 | mid_secretory |
| GSM742069 | 4 | 0 | TRAIN179 | proliferative |
| GSM742070 | 4 | 0 | TRAIN179 | proliferative |
| GSM742071 | 4 | 0 | TRAIN179 | early_secretory |
| GSM742072 | 4 | 0 | TRAIN179 | proliferative |
| GSM742073 | 4 | 0 | TRAIN179 | early_secretory |
| GSM742074 | 4 | 0 | TRAIN179 | proliferative |
| GSM742075 | 4 | 0 | TRAIN179 | mid_secretory |
| GSM742076 | 4 | 0 | TRAIN179 | proliferative |
| GSM742077 | 4 | 0 | TRAIN179 | early_secretory |
| GSM742078 | 4 | 0 | TRAIN179 | mid_secretory |
| GSM742079 | 4 | 0 | TRAIN179 | proliferative |
| GSM742080 | 4 | 0 | TRAIN179 | early_secretory |

#### Supplementary Table S3. Results of 37 individual test-label changes

| GSM | Truth | Supplied | Before | After | Shift (SD) | Switch |
|---|---|---|---|---|---|---|
| GSM150190 | 1 | 0 | -0.597 | -3.752 | 0.750 | 0 |
| GSM150191 | 1 | 0 | +8.659 | +4.922 | 0.888 | 0 |
| GSM150192 | 1 | 0 | -0.985 | -4.194 | 0.762 | 0 |
| GSM150193 | 1 | 0 | +3.832 | +0.524 | 0.786 | 0 |
| GSM150194 | 1 | 0 | +2.755 | -0.463 | 0.764 | 1 |
| GSM150195 | 1 | 0 | +2.810 | -0.347 | 0.750 | 1 |
| GSM150196 | 0 | 1 | -3.368 | -0.084 | 0.780 | 0 |
| GSM150197 | 0 | 1 | -4.008 | -0.777 | 0.767 | 0 |
| GSM150198 | 0 | 1 | -2.259 | +0.913 | 0.753 | 1 |
| GSM150199 | 0 | 1 | -3.788 | -0.547 | 0.770 | 0 |
| GSM150201 | 0 | 1 | -4.175 | -1.029 | 0.747 | 0 |
| GSM150202 | 1 | 0 | +2.163 | -1.031 | 0.759 | 1 |
| GSM150203 | 1 | 0 | +2.595 | -0.592 | 0.757 | 1 |
| GSM150204 | 1 | 0 | +3.845 | +0.548 | 0.783 | 0 |
| GSM150205 | 1 | 0 | +2.089 | -1.140 | 0.767 | 1 |
| GSM150206 | 1 | 0 | +8.916 | +5.304 | 0.858 | 0 |
| GSM150207 | 1 | 0 | +2.806 | -0.472 | 0.779 | 1 |
| GSM150208 | 0 | 1 | -1.622 | +1.608 | 0.767 | 1 |
| GSM150209 | 0 | 1 | -3.391 | -0.121 | 0.777 | 0 |
| GSM150210 | 0 | 1 | -0.307 | +2.885 | 0.758 | 1 |
| GSM150211 | 1 | 0 | +1.228 | -1.929 | 0.750 | 1 |
| GSM150212 | 1 | 0 | +4.868 | +1.531 | 0.793 | 0 |
| GSM150213 | 1 | 0 | +6.670 | +3.228 | 0.818 | 0 |
| GSM150214 | 1 | 0 | +9.671 | +5.876 | 0.901 | 0 |
| GSM150215 | 1 | 0 | +6.438 | +3.109 | 0.791 | 0 |
| GSM150216 | 1 | 0 | +8.314 | +4.644 | 0.872 | 0 |
| GSM150217 | 1 | 0 | +1.687 | -1.501 | 0.757 | 1 |
| GSM150218 | 1 | 0 | +2.317 | -0.885 | 0.761 | 1 |
| GSM150219 | 1 | 0 | +3.330 | +0.124 | 0.762 | 0 |
| GSM150220 | 0 | 1 | -4.370 | -1.109 | 0.775 | 0 |
| GSM150221 | 0 | 1 | -1.767 | +1.400 | 0.752 | 1 |
| GSM150222 | 0 | 1 | -3.664 | -0.417 | 0.771 | 0 |
| GSM150223 | 0 | 1 | -3.985 | -0.698 | 0.781 | 0 |
| GSM150224 | 0 | 1 | -1.904 | +1.281 | 0.757 | 1 |
| GSM150225 | 0 | 1 | -2.890 | +0.283 | 0.754 | 1 |
| GSM150226 | 0 | 1 | -2.027 | +1.165 | 0.758 | 1 |
| GSM150227 | 0 | 1 | -3.262 | -0.054 | 0.762 | 0 |

### Supplementary Table S4. Original and permuted-training-label accuracy in 18 conditions

| Layout | Strata | Process | Model | BA | Median | Q05 | Q95 | All-control |
|---|---|---|---|---|---|---|---|---|
| FULL179 | BATCH | NO OUTCOME | ARCHIVED | 0.786 | 0.551 | 0.370 | 0.731 | 0 |
| FULL179 | BATCH | NO OUTCOME | SENSITIVITY | 0.754 | 0.534 | 0.361 | 0.723 | 0 |
| FULL179 | BATCH | OUTCOME | ARCHIVED | 0.952 | 0.850 | 0.701 | 0.929 | 0 |
| FULL179 | BATCH | OUTCOME | SENSITIVITY | 0.952 | 0.835 | 0.676 | 0.930 | 0 |
| FULL179 | BATCH PHASE | NO OUTCOME | ARCHIVED | 0.786 | 0.558 | 0.362 | 0.748 | 0 |
| FULL179 | BATCH PHASE | NO OUTCOME | SENSITIVITY | 0.754 | 0.565 | 0.344 | 0.740 | 0 |
| FULL179 | BATCH PHASE | OUTCOME | ARCHIVED | 0.952 | 0.857 | 0.707 | 0.952 | 0 |
| FULL179 | BATCH PHASE | OUTCOME | SENSITIVITY | 0.952 | 0.842 | 0.691 | 0.952 | 0 |
| DEDUP153 | BATCH | NO OUTCOME | ARCHIVED | 0.659 | 0.549 | 0.361 | 0.714 | 0 |
| DEDUP153 | BATCH | NO OUTCOME | SENSITIVITY | 0.604 | 0.542 | 0.354 | 0.714 | 0 |
| DEDUP153 | BATCH | OUTCOME | ARCHIVED | 0.952 | 0.881 | 0.778 | 0.952 | 0 |
| DEDUP153 | BATCH | OUTCOME | SENSITIVITY | 0.976 | 0.857 | 0.707 | 0.952 | 0 |
| DEDUP153 | BATCH | RMA ONLY | ARCHIVED | 0.500 | 0.500 | 0.500 | 0.500 | 191 |
| DEDUP153 | BATCH | RMA ONLY | SENSITIVITY | 0.500 | 0.500 | 0.500 | 0.548 | 163 |
| FULL179 | BATCH | RMA ONLY | ARCHIVED | 0.619 | 0.500 | 0.500 | 0.524 | 165 |
| FULL179 | BATCH | RMA ONLY | SENSITIVITY | 0.595 | 0.500 | 0.500 | 0.548 | 142 |
| FULL179 | BATCH PHASE | RMA ONLY | ARCHIVED | 0.619 | 0.500 | 0.500 | 0.524 | 176 |
| FULL179 | BATCH PHASE | RMA ONLY | SENSITIVITY | 0.595 | 0.500 | 0.500 | 0.595 | 151 |

#### Supplementary Table S5. Balanced-accuracy differences for matched ComBat refits

| Layout | Strata | Model | Median | Q05 | Q95 | Positive |
|---|---|---|---|---|---|---|
| FULL179 | BATCH | ARCHIVED | +0.284 | +0.150 | +0.426 | 199/199 |
| FULL179 | BATCH | SENSITIVITY | +0.284 | +0.118 | +0.442 | 199/199 |
| FULL179 | BATCH PHASE | ARCHIVED | +0.269 | +0.095 | +0.449 | 199/199 |
| FULL179 | BATCH PHASE | SENSITIVITY | +0.260 | +0.102 | +0.457 | 198/199 |
| DEDUP153 | BATCH | ARCHIVED | +0.317 | +0.174 | +0.498 | 199/199 |
| DEDUP153 | BATCH | SENSITIVITY | +0.317 | +0.157 | +0.473 | 199/199 |

#### Supplementary Table S6. Complete results for 1,194 matched comparisons

The full-precision results are available in the machine-readable file `supplementary_data/Supplementary_Table_S6_1194_paired_deltas.csv`. For each comparison, it records `layout`, `family`, `configuration`, `replicate`, and `delta_BA`. All 1,194 matched comparisons are included in the supplementary data without omissions.

### S7. A shared source cohort can contain very different kinds of evidence

In the deduplicated 153-array training roster, the source and outcome breakdown was GSE51981: 56 ENDO and 62 controls; GSE29981: 20 controls only; GSE4888: 15 controls only. GSE29981 profiles a glandular epithelial preparation, unlike the bulk endometrium that makes up the principal GSE51981 analysis. These differences can affect intensity levels before any classifier is fitted. Across 324 fixed probe IDs, the median absolute ENDO–control contrast within GSE51981 was 0.4104 $\log_2$ units; source-control differences against the GSE51981 controls were 2.0277 for GSE29981, 0.7233 for GSE4888 and 1.1282 for the 35 combined additional controls. The corresponding proportions of probes with a larger between-control-source than within-source diagnosis contrast were 89.5%, 67.9% and 77.5%. Probe IDs are neither mutually independent nor automatically marker genes.

The three uncombined phase groups within GSE51981 each contained both ENDO and controls: early secretory 13/12, mid-secretory 19/18 and proliferative 24/32 in the frozen training subset. Only 164/324 probes had effects of the same sign across all three phases. These are descriptive measurements, not significant, independently replicated biomarkers.

Table 6: GSE51981 scanner-year composition and exact overlap with GSE6364 (source metadata).

| Year | ENDO | OTHER | NUPP | Exact CEL copies |
|---|---|---|---|---|
| 2004 | 11 | 0 | 6 | 17 |
| 2005 | 8 | 0 | 1 | 9 |
| 2009 | 46 | 31 | 0 | 0 |
| 2010 | 2 | 0 | 15 | 0 |
| 2011 | 10 | 6 | 12 | 0 |

### S8. Fold-contained internal benchmark and acquisition-date negative controls

All 148 GSE51981 source CELs were processed independently by MAS5 (118 archived reference vectors plus 30 new ones). The fixed 193-gene/324-probe representation is reproducibly derivable from a 275-gene core list and GPL570/GPL571 annotation overlaps. The existence of such a label-free construction does not on its own certify the historical chronological choice of the feature rule. The experiment used five outer folds, four inner folds, $C \in \{0.01, 0.1, 1, 10\}$, and train-fold scaling. Donor IDs were not authenticated; this is sample-level exploratory out-of-fold evaluation.

Table 7: Task definitions and sample-level evaluation metrics.

| Task | ENDO | Controls | ROC AUC | Balanced acc. |
|---|---|---|---|---|
| ENDO vs all non-ENDO | 77 | 71 | .7556 | .7017 |
| ENDO vs OTHER | 77 | 37 | .7329 | .6430 |
| ENDO vs NUPP | 77 | 34 | .8934 | .8031 |
| 2009, ENDO vs OTHER | 46 | 31 | .7237 | .6220 |

A post-hoc scanner-date-only negative control obtained out-of-fold AUC 0.7365 for ENDO/all, 0.6114 for ENDO/OTHER, 0.9347 for ENDO/NUPP and 0.5361 for 2009 ENDO/OTHER. Within-date pairwise rank concordance for fixed molecular predictions was 0.6850 (400 pairs), 0.6450 (338), 0.6774 (62) and 0.7000 (290), respectively. The within-date quantity is not a retrained within-date diagnostic AUC.

### S9. The post-hoc 2009 held-out-date experiment

The 2009 cohort had 46 ENDO and 31 OTHER arrays across five eligible acquisition dates. When each date was withheld from both model training and inner selection, the observed date-specific AUCs were:

| Date held out | ENDO | OTHER | AUC |
|---|---|---|---|
| 8 Jan 2009 | 3 | 5 | .5333 |
| 14 Jan 2009 | 10 | 11 | .7182 |
| 15 Jan 2009 | 17 | 5 | .7765 |
| 22 Jan 2009 | 8 | 8 | .7344 |
| 6 Feb 2009 | 8 | 2 | .6250 |

Pooled ROC AUC was 0.698; the unweighted mean of the five date-specific AUCs was 0.678. The endpoints weight scanner dates differently. The 199 date-stratified training-label-permutation add-one fractions were 0.030 and 0.070, respectively. This is a post-hoc analysis on one cohort with sparse held-out dates, not a prospective validation. The full date-specific results are also included in the supplementary data.

#### S10. Five-gene Huang-inspired RF reconstruction

The fixed genes were ACKR1, LMNB1, MFAP4, NMU and SEMA3C. GSE6364 (37 CEL) plus GSE25628 (22 CEL) supplied 59 training profiles. Five independently processed gene signals and a 1,000-tree scikit-learn forest define a *new reconstruction*; the original author-trained model and original `sva` transformation were not recovered. Exactly 26 of the GSE51981 test arrays had byte-identical GSE6364 training counterparts. Every own-copy omission retained 58 training samples, as did the same-class control omission. The own-copy-out AUC values from five fixed seeds were .3233, .3045, .3083, .2932 and .3233. The class-stratified 5,000-resample 95% conditional interval for the base-seed own-copy-out AUC was .1053–.5714, and that for the baseline perfect ranking was [1,1], conditional on resampling the fixed observed predictions. The former interval includes .5. These numbers do not correct the authors' published AUC .656 or measure clinical performance on independent patients.

Table 8: AUC under exact training-copy omission. The last two rows pool scores from different models.

| Fixed test set | Copy present | Own copy omitted | Other same-class omitted |
|---|---|---|---|
| 26 repeated CELs | 1.0000 | .3233 | 1.0000 |
| 111 ENDO/NUPP (hybrid) | .8657 | .7086 | .8641 |
| 148 ENDO/non-ENDO (hybrid) | .7545 | .6116 | .7533 |

#### S11. Published marker panel and calendar overlap support

We assessed the feasibility of comparing ten previously published genes reported by Huang et al. (2022) (ACKR1, LMNB1, MFAP4, NMU and SEMA3C) and Chen et al. (2023) (CHPF, PDK3, CITED2, GPC3 and ADH6) within common scanner dates in GSE51981. The ten genes were measurable through 19 GPL570 probe identifiers and individually processed MAS5 profiles. The two disease comparisons had markedly different calendar support (Table 9).

Table 9: Observed sample support on scanner dates with both classes; this is a feasibility table, not a diagnostic accuracy estimate.

| Comparison | Eligible CEL arrays | Shared scanner dates |
|---|---|---|
| ENDO versus OTHER | 91 | 6 |
| ENDO versus NUPP | 42 | 4 |

Table S11: Published-marker ENDO–control $\log_2$ contrasts before and after restricting to common scanner dates (20 fixed gene–task tests). Intervals are sample-bootstrap 95% CIs. The last two effect columns use identical arrays; FE denotes scanner-date fixed effects. P values are descriptive label-shuffle tests (full and matched) or within-date permutations (FE). Holm adjustment applies to the 20 FE tests.

| Task | Gene | All dates, naive (95% CI) | Matched dates, naive (95% CI) | Same matched dates, date FE (95% CI) | $p_{\mathrm{full}}$ | $p_{\mathrm{matched}}$ | $p_{\mathrm{FE}}$ | Holm $p$ |
|---|---|---|---|---|---|---|---|---|
| ENDO_vs_OTHER | ACKR1 | +0.076 [-0.813, +0.975] | +0.715 [-0.063, +1.417] | +0.862 [+0.118, +1.586] | 0.878 | 0.138 | 0.043 | 0.602 |
| ENDO_vs_OTHER | LMNB1 | -0.187 [-0.526, +0.148] | -0.232 [-0.534, +0.029] | -0.363 [-0.671, -0.097] | 0.289 | 0.177 | 0.029 | 0.440 |
| ENDO_vs_OTHER | MFAP4 | -0.271 [-1.002, +0.376] | +0.289 [-0.291, +0.898] | +0.226 [-0.329, +0.856] | 0.442 | 0.358 | 0.491 | 1.000 |
| ENDO_vs_OTHER | NMU | +0.101 [-0.464, +0.707] | +0.320 [-0.215, +0.917] | +0.088 [-0.450, +0.661] | 0.714 | 0.276 | 0.748 | 1.000 |
| ENDO_vs_OTHER | SEMA3C | +0.146 [-0.165, +0.440] | -0.145 [-0.429, +0.132] | -0.214 [-0.517, +0.068] | 0.427 | 0.394 | 0.203 | 1.000 |
| ENDO_vs_OTHER | CHPF | +0.270 [+0.037, +0.511] | +0.279 [+0.066, +0.499] | +0.305 [+0.072, +0.544] | 0.051 | 0.059 | 0.028 | 0.440 |
| ENDO_vs_OTHER | PDK3 | +0.206 [-0.035, +0.452] | -0.186 [-0.395, +0.020] | -0.171 [-0.393, +0.051] | 0.174 | 0.125 | 0.171 | 1.000 |
| ENDO_vs_OTHER | CITED2 | +0.034 [-0.372, +0.460] | +0.111 [-0.333, +0.571] | +0.250 [-0.183, +0.660] | 0.881 | 0.660 | 0.327 | 1.000 |
| ENDO_vs_OTHER | GPC3 | +0.593 [+0.279, +0.905] | +0.380 [+0.075, +0.687] | +0.405 [+0.096, +0.739] | 0.002 | 0.019 | 0.017 | 0.306 |
| ENDO_vs_OTHER | ADH6 | +0.409 [+0.063, +0.762] | +0.219 [-0.160, +0.556] | +0.155 [-0.231, +0.523] | 0.037 | 0.261 | 0.444 | 1.000 |
| ENDO_vs_NUPP | ACKR1 | +2.347 [+1.478, +3.308] | +1.121 [-0.088, +2.362] | +1.173 [-0.639, +3.047] | 0.001 | 0.148 | 0.228 | 1.000 |
| ENDO_vs_NUPP | LMNB1 | -1.174 [-1.642, -0.684] | -1.781 [-2.320, -1.286] | -1.388 [-1.835, -0.933] | 0.001 | 0.001 | 0.002 | 0.030 |
| ENDO_vs_NUPP | MFAP4 | +0.727 [+0.025, +1.433] | -0.093 [-1.016, +0.763] | -0.821 [-1.688, +0.029] | 0.036 | 0.868 | 0.257 | 1.000 |
| ENDO_vs_NUPP | NMU | -0.396 [-0.987, +0.184] | -1.109 [-1.604, -0.548] | +0.298 [-0.125, +0.742] | 0.197 | 0.009 | 0.551 | 1.000 |
| ENDO_vs_NUPP | SEMA3C | -0.629 [-0.961, -0.293] | -0.470 [-0.962, -0.002] | -0.133 [-0.767, +0.471] | 0.001 | 0.105 | 0.742 | 1.000 |
| ENDO_vs_NUPP | CHPF | +1.163 [+0.820, +1.453] | +0.943 [+0.705, +1.219] | +0.672 [+0.349, +1.011] | 0.001 | 0.001 | 0.005 | 0.095 |
| ENDO_vs_NUPP | PDK3 | -0.159 [-0.421, +0.110] | -0.235 [-0.417, -0.057] | -0.157 [-0.402, +0.081] | 0.303 | 0.201 | 0.309 | 1.000 |
| ENDO_vs_NUPP | CITED2 | +1.450 [+0.990, +1.891] | +1.431 [+0.820, +2.015] | +0.999 [+0.265, +1.718] | 0.001 | 0.001 | 0.020 | 0.340 |
| ENDO_vs_NUPP | GPC3 | +0.508 [+0.091, +0.901] | +0.740 [+0.268, +1.187] | +0.284 [-0.230, +0.752] | 0.011 | 0.028 | 0.417 | 1.000 |
| ENDO_vs_NUPP | ADH6 | -0.023 [-0.355, +0.321] | -0.078 [-0.585, +0.446] | +0.026 [-0.572, +0.625] | 0.905 | 0.807 | 0.949 | 1.000 |

For each of the 20 fixed gene–comparison combinations, Supplementary Table S11 reports three complementary estimates: (i) an unadjusted ENDO–control $\log_2$ contrast on all eligible arrays, (ii) an unadjusted contrast restricted to scanner dates containing both clinical groups, and (iii) a scanner-date fixed-effect contrast on *exactly the same restricted arrays* as (ii). The first-to-second difference therefore reflects restriction to shared-date support, whereas the second-to-third difference describes adjustment within that support. These changes should not be interpreted as causal contributions of acquisition date. For ENDO/OTHER, 54 ENDO and 37 OTHER arrays share six dates (91 total); for ENDO/NUPP, 14 ENDO and 28 NUPP arrays share four dates (42 total). Restriction leaves substantial uncertainty for the latter comparison.

The decomposition is informative. For ACKR1 in ENDO/NUPP, the unadjusted contrast decreased from +2.347 $\log_2$ across all arrays to +1.121 on common dates, but was +1.173 after date adjustment *on those same arrays*. Thus, most of the apparent reduction in this example is associated with changing which samples are eligible, not the fitted date covariate. For PDK3 in ENDO/OTHER, the sign changed from +0.206 (all arrays) to $-0.186$ (common-date arrays) and remained $-0.171$ after date adjustment. Other markers showed different combinations of support and adjustment effects; they were not selected based on their results.

Only one of the 20 scanner-date fixed-effect tests met Holm-adjusted $p < 0.05$: LMNB1 for ENDO versus NUPP ($-1.388$ $\log_2$, 95% sample-bootstrap CI $[-1.835, -0.933]$, adjusted $p = 0.030$). The unadjusted permutation comparisons are descriptive and do not preserve date strata; the fixed-effect permutation tests reassign labels within date, and Holm adjustment was applied to the 20 fixed-effect tests. All confidence intervals use sample-level rather than verified-donor-level bootstrap resampling. This post-hoc examination of published genes is a sensitivity analysis, not independent biomarker validation or an attempt to reconstruct either published multigene classifier.

### S12. Literature screen, reporting checklist, and source files

A targeted Europe PMC search produced 246 bibliographic records; 29 returned indexed hits for both series and were individually classified, with full methods available for 27. Multiple use patterns were present. The cross-study table in the main text reports the strongest documentary evidence. An inclusion count inferred from published totals remains *conditional* without a sample-level source roster, even when the underlying GEO datasets share exact CEL files. The accompanying machine-readable literature-screening table lists retained and excluded candidates, original study references and evidence grades.

**Suggested GEO-LEAK reporting checks.** (1) Hash decompressed raw measurements and build exact duplicate clusters; (2) map donors and specimens separately from digital file identity; (3) verify platform, preparation, tissue and phase compatibility; (4) compare acquisition dates and cohort source against outcome labels; (5) prevent held-out outcomes from entering normalization, feature development or calibration; (6) specify a realistic procedure for entirely unseen batches; and (7) publish the split manifest, null findings and explicit unknowns. This is a proposed research checklist, not an endorsed clinical standard.

Supplementary data provide the original full-precision Tables S1–S6, source manifests and selected score summaries. The accompanying research-preview code includes figure-generation scripts. The audit does not certify donor independence or prospective clinical validity, and raw CEL archives are not redistributed.

### S13. Additional information-flow and intervention figures

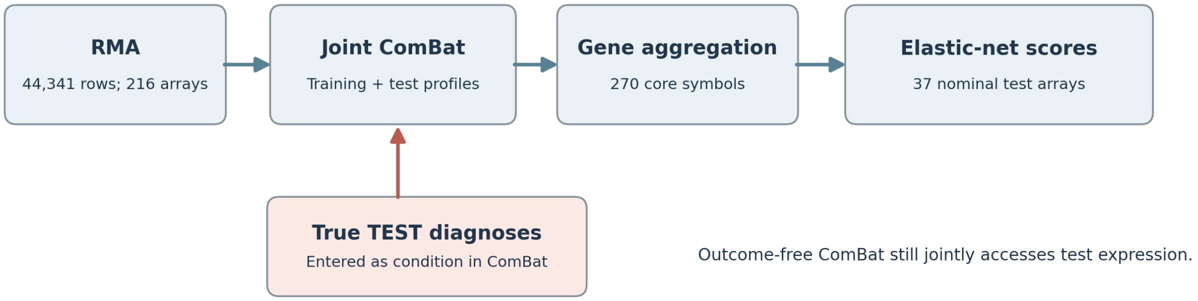


Figure 1: **Supplementary Figure S1.** Where unknown test outcomes entered the reconstructed joint ComBat design. This illustration documents the audited code's information path rather than a universal property of ComBat.

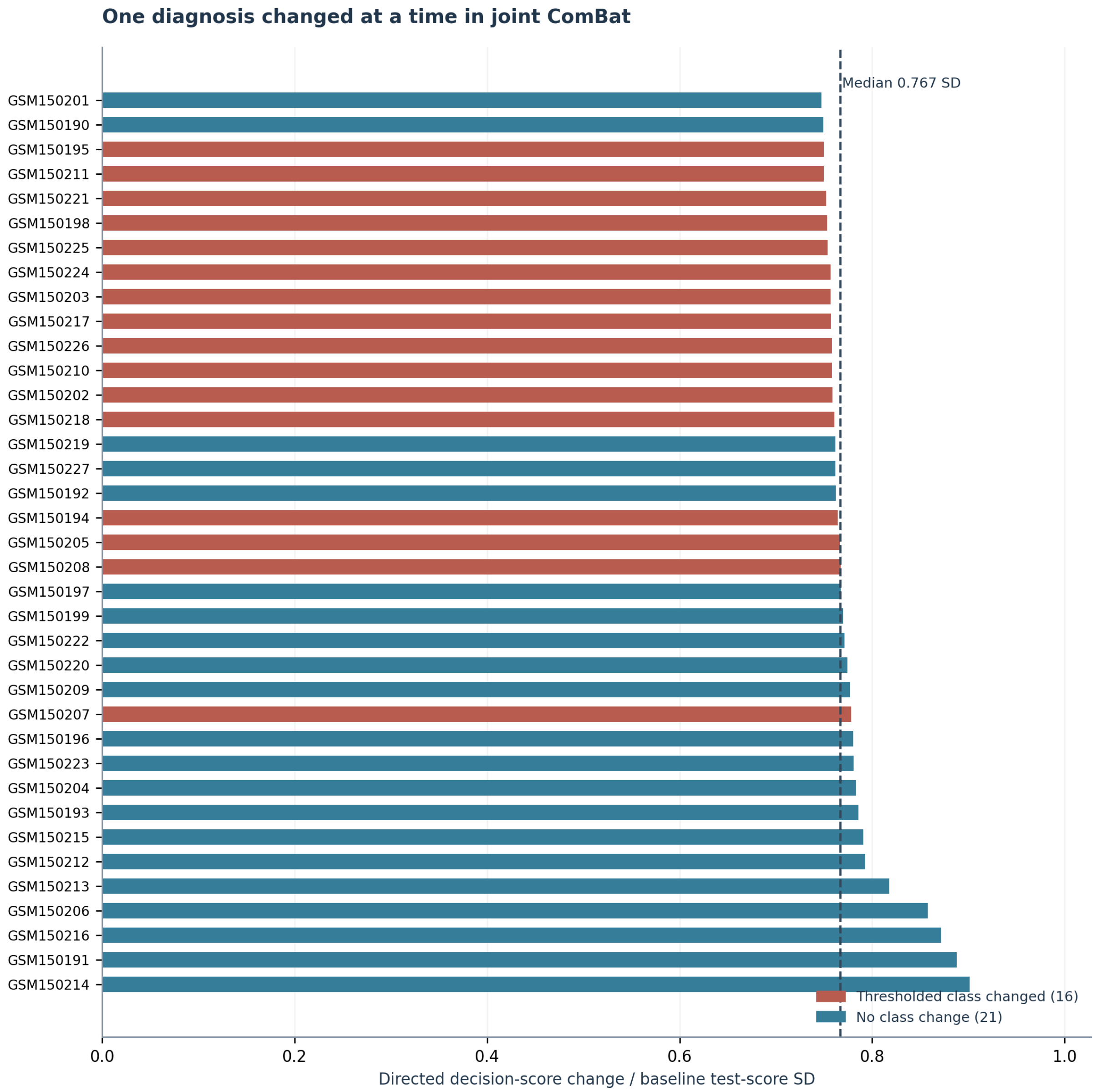


Figure 2: **Supplementary Figure S2.** Test-label interventions under a fixed classifier. Each intervention changed one diagnosis supplied to ComBat, not the expression measurement or the true scoring outcome.

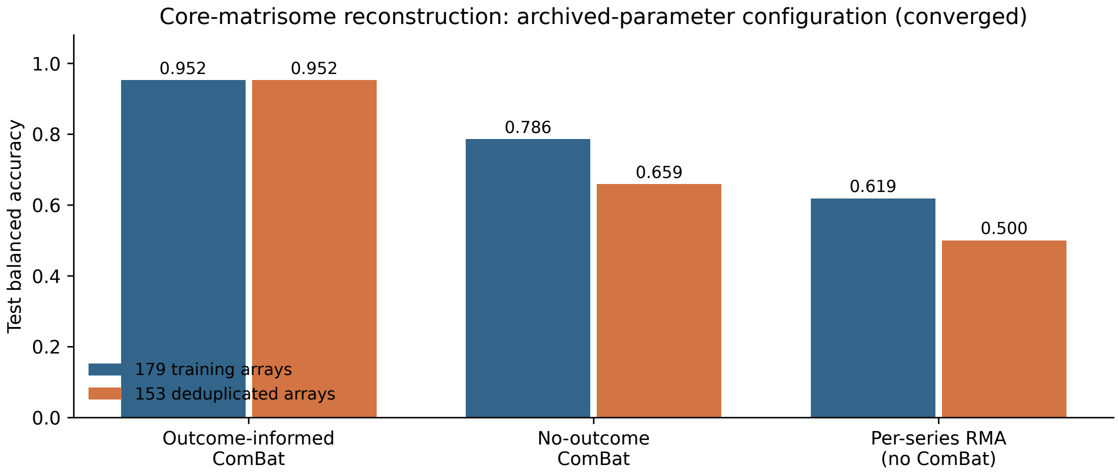


Figure 3: **Supplementary Figure S3.** Converged factorial comparison of original and deduplicated training layouts under three preprocessing arms. Both joint ComBat arms use the held-out expression distribution; one additionally uses true held-out diagnoses.
